# Simulation-Based Study on the COVID-19 Airborne Transmission in a Restaurant

**DOI:** 10.1101/2020.12.10.20247403

**Authors:** Han Liu, Sida He, Lian Shen, Jiarong Hong

## Abstract

COVID-19 has shown a high potential of transmission via virus-carrying aerosols as supported by growing evidence. However, detailed investigations that draw direct links between aerosol transport and virus infection are still lacking. To fill in the gap, we conducted a systematic computational fluid dynamics (CFD)-based investigation of indoor air flow and the associated aerosol transport in a restaurant setting, where likely cases of airborne infection of COVID-19 caused by asymptomatic individuals were widely reported by the media. We employed an advanced in-house large eddy simulation (LES) solver and other cutting-edge numerical methods to resolve complex indoor processes simultaneously, including turbulence, flow–aerosol interplay, thermal effect, and the filtration effect by air conditioners. Using the aerosol exposure index derived from the simulation, we are able to provide a spatial map of the airborne infection risk under different settings. Our results have shown a remarkable direct linkage between regions of high aerosol exposure index and the reported infection patterns in the restaurant, providing strong support to the airborne transmission occurring in this widely-reported incidence. Using flow structure analysis and reverse-time tracing of aerosol trajectories, we are able to further pinpoint the influence of environmental parameters on the infection risks and highlight the needs for more effective preventive measures, e.g., placement of shielding according to the local flow patterns. Our research, thus, has demonstrated the capability and value of high-fidelity CFD tools for airborne infection risk assessment and the development of effective preventive measures.

## I. INTRODUCTION

The COVID-19 pandemic (caused by the SARS-CoV-2) has led to more than 69 million infections and 1.5 million deaths as of writing. A growing number of evidence has suggested airborne transmission of respiratory droplets, either both produced from asymptomatic or pre-symptomatic individuals or none, is a potential pathway that contributes to the wide spread of the disease^1–4^. In particular, an incidence in a restaurant in Guangzhou, China, where a single asymptomatic COVID-19 patient caused the infection of eight people sitting in the same and two adjacent tables, has been widely reported in the media^5,6^ and recent scientific literature^1^ as a hallmark demonstration of such transmission pathway. Specifically, for airborne transmission, viruses attached to small respiratory droplets (typically < 5 *µ*m, referred to as aerosols hereafter) produced by even normal breathing or talking can stay suspended in the air for hours, and they can accumulate, particularly in indoor spaces with poor ventilation, imposing high infection risk to individuals who inhale them^7–11^. Under such circumstances, the commonly adopted social distancing rules can no longer serve as a proper preventive measure^12^. As a result, there is a dire need for science-driven risk assessment tools that can provide actionable and more appropriate guidelines under different practical settings. Despite numerous analytical and numerical modeling work that provide the first order estimate of aerosol exposure and corresponding airborne infection risk^13–15^, only recently, the computational fluid dynamics (CFD) tools have been actively employed to provide a more precise risk assessment for aerosol exposure, especially, its spatial and temporal variations in indoor spaces. Specifically, using Reynolds-averaged Navier-Stokes (RANS) simulation with Lagrangian particle models, Shao *et al*. (2020)^16^ investigated the airborne transmission risk associated with an asymptomatic patient is representative elevator, classroom, and grocery store settings. They introduced a local risk index and evaluated its spatial variation under different ventilation settings, which reveals indoor infection hot spots (corresponding to high risk of aerosol exposure) due to inappropriate ventilation design. Mathai *et al*. (2020)^17^ uses RANS to explore how the ventilation configuration of open and closed windows influences the airborne transmission in a passenger car cabin. The aerosols are modeled as passive scalars by an advection-diffusion equation. Subsequently, Vuorinen *et al*. (2020)^18^ performed large-eddy simulations (LES) and Monte-Carlo simulations to understand the exposure risk of the moving people in a public indoor environment where an infected person coughs. They showed that the exposure time for getting infected can range from seconds to hours depending on the aerosol diameter and indoor turbulence level, and the spatial extent of high-risk zone can be as large as 4 m from the infected person. Remarkably, Li *et al*. (2020)^9^ conducted a simulation of air flows in the aforementioned restaurant case and used the results to infer the strong influence of air conditioning and ventilation on the reported breakout. In spite of these vigorous efforts, there have been very few CFD studies so far that can offer direct linkage between the simulation results and reported infection patterns as well as the corresponding detailed physical mechanisms that lead to airborne disease transmission. Such work is crucial not only to showcase the validity of CFD to capture airborne transmission processes, but also help us develop pinpointed interventions for mitigating the disease spread.

To fill in the gap, in this paper we present a systematic LES based investigation of indoor airflow and the associated aerosol transport in the aforementioned restaurant setting, where likely cases of airborne infection caused by asymptomatic individuals were reported and the detailed information of infection process through contact tracing are available. Our simulation employs an advanced in-house CFD solver and uses cutting-edge numerical methods (i.e., advanced immersed boundary (IB) method, stochastic modeling of Brownian motion and effect of subgrid-scale (SGS) flow structures on aerosols, Cunningham slip correction, etc.) to resolve complex indoor processes, including turbulence, flow– aerosol interaction, thermal effect, and the filtration effect by air conditioners. Such a combination of high-fidelity numerical methods, several of which has not been adopted in the past simulation work of indoor airflow, allows us to depict a detailed picture of airborne transmission in this hallmark incidence and derive a series of important insights that can benefit our understanding of this transmission pathway and the development of more effective preventive measures. The rest of this paper is organized as follows. The methodology used in our simulation is described in Sec. II. Subsequently, we provide the results of simulation under different ventilation and thermal settings in conjunction with their potential connection with reported infection in Sec. III, with conclusions given in Sec. IV.

## II. NUMERICAL METHOD AND SIMULATION SET-UP

### A. Numerical method

In this study, LES is conducted, where the SGS stress is modeled by the dynamic Smagorinsky model^19–21^. The governing equations are

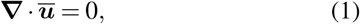

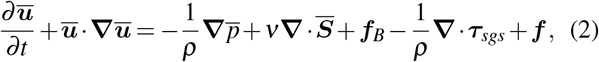

where ***u*** is the velocity vector, *ρ* is the density, *p* is the pressure, *v* is the kinematic viscosity, ***S*** = (**∇*u*** + **∇*u***^*T*^)*/*2 is the strain rate tensor, ***f***_*B*_ is the buoyancy force, ***τ***_*sgs*_ is the SGS stress tensor, ***f*** represents the forces exerted by the solid structures inside the room in the IB method, and the overbar denotes the spatial filter in LES. In the case where the turbulence in the inner boundary layer on the walls is not fully resolved, a log-law-based wall model^22^ is used. The secondorder central differencing scheme is used for the spatial discretization of both the convection and diffusion terms, and the second-order Runge–Kutta method is used for time integration. The equations are solved by the fractional-step method, and the discretized Poisson equation for pressure is solved by the Portable, Extensible Toolkit for Scientific Computation (PETSc)^23^.

Thermal effect is accounted for in the simulation, due to the consideration that the human bodies around tables and the hot foods on the tables can lead to temperature variations, which create thermal plumes in the room. Heat transfer is simulated by an advection–diffusion equation for temperature as,

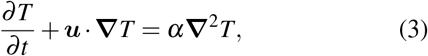

where *T* denotes the fluid temperature and *α* = 22.39 *×* 10^−6^ m^2^/s is the thermal diffusivity. To couple the thermal effect with the airflow dynamics, the Boussinesq approximation^24^ is applied to Eq. (2):

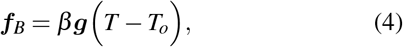

where ***g*** is the gravitational acceleration, *β* = 3400 *×* 10^−6^ /°C is the coefficient of thermal expansion and *T*_*o*_ is the reference temperature.

The influence of the solid structures on the flow dynamics is simulated by the IB method^25–29^. This method can be used on a static Cartesian grid to comply with the structure geometry, and is thus convenient to apply to structures with complex geometry. In the IB method, the velocity boundary condition is satisfied by interpolating the velocity to the grid points adjacent to the structure surfaces according to the velocity profile of the boundary layer. Linear and nonlinear interpolations are applied for cases where the boundary layer is resolved and modeled, respectively.

The transport of aerosols is simulated with a Lagrangian particle tracking algorithm. The aerosol location ***x***_*p*_ is solved by

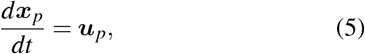

where ***u***_*p*_ is the aerosol velocity obtained by

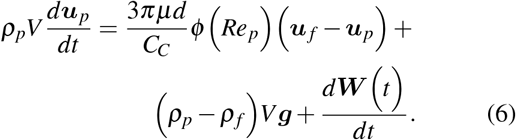

In the above equation, the first and second terms on the right-hand side represent the Stokes drag force and gravity/buoyancy force, where *ρ*_*p*_ and *ρ*_*f*_ are the density of the aerosols and the fluid, respectively, ***u***_*p*_ is the aerosol velocity, ***u*** _*f*_ is the velocity of the fluid at the aerosol location, *d* is the aerosol diameter, 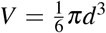 is the aerosol volume, *µ* is the fluid dynamic viscosity, *Re*_*p*_ = *ρ*_*f*_ *d*|***u*** _*f*_ − ***u***_*p*_|*/µ* is the aerosol Reynolds number, 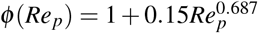, and *C*_*C*_ is the Cunningham correction factor^30^. Note that the Cunningham correction is applied to the Stokes drag force, because the aerosols are so small that the surrounding gas cannot be modeled as a continuum medium and the non-slip boundary condition on the aerosol surface cannot be strictly applied^31–33^. The third term of the right-hand side represents the Brownian motion effect, where each component of ***W*** (*t*) is a Wiener process with zero mean and variance 2*γk*_*B*_*T* with 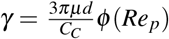 and *k*_*B*_ the Boltzmann constant^34,35^. Both equations (5) and (6) are numerically solved by an Adam– Bashforth method. To capture the aerosol deposition effect, an aerosol whose center has a distance from the wall less than its radius is immediately removed from the computation domain.

The effect of the SGS flows on the aerosol dynamics is accounted for by a stochastic model^36,37^. In this model, the fluid velocity at the aerosol location ***u***_*f*_ is decomposed into a grid-resolvable part ***u***_*f,r*_ and a SGS part ***u***_*f,s*_,

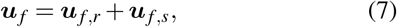

where ***u***_*f,r*_ is obtained by interpolating the flow velocity values on the grid nodes to the aerosol location by a fourth-order interpolation scheme and ***u***_*f,s*_ is modeled by the following Langevin equation,

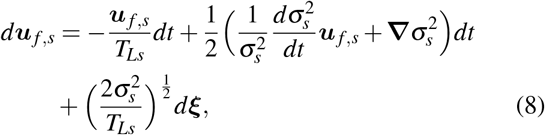

where 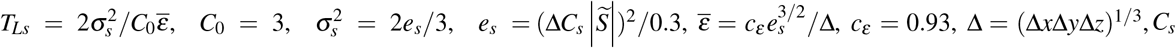 is the Smagorinsky coefficient, 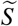 is the filtered rate-of-strain tensor, and each components of *d****ξ*** is a Gaussian random variable with zero mean and variance *dt*. Equation (8) is integrated in time with an implicit algorithm for numerical stability.

The aerosol volume fraction in the present study is *ϕ* = 4 *×* 10^−7^, which is much smaller than 10^−3^, the threshold of fourway coupling^30^. Therefore, the aerosol-aerosol interaction is not considered.

Our numerical tools are validated by a forced convection case and a mixed convection case. See Appendix A for the validation details.

### B. Background of study and simulation set-up

The present numerical study is based on a real infection event that occurred in a restaurant in Guangzhou, China, which is illustrated in Fig. 1. On January 24^*th*^, 2020, there were 89 customers dining in the restaurant during the lunch time. After this lunch, nine people sitting on tables number one, two, and three [labeled in Fig. 1(a) as T1, T2, and T3] were infected. The other people sitting away from these three tables were not infected^9^. It has been believed that three people eating at T1 and two people at T3 were infected directly during this lunch^1^. Virus was transmitted from patient zero sitting at T2. In order to simplify the discussion, customers are labeled as TAPBs as shown in Fig. 2, where A stands for the index of the tables and B stands for the index of the customers.

**FIG. 1.**
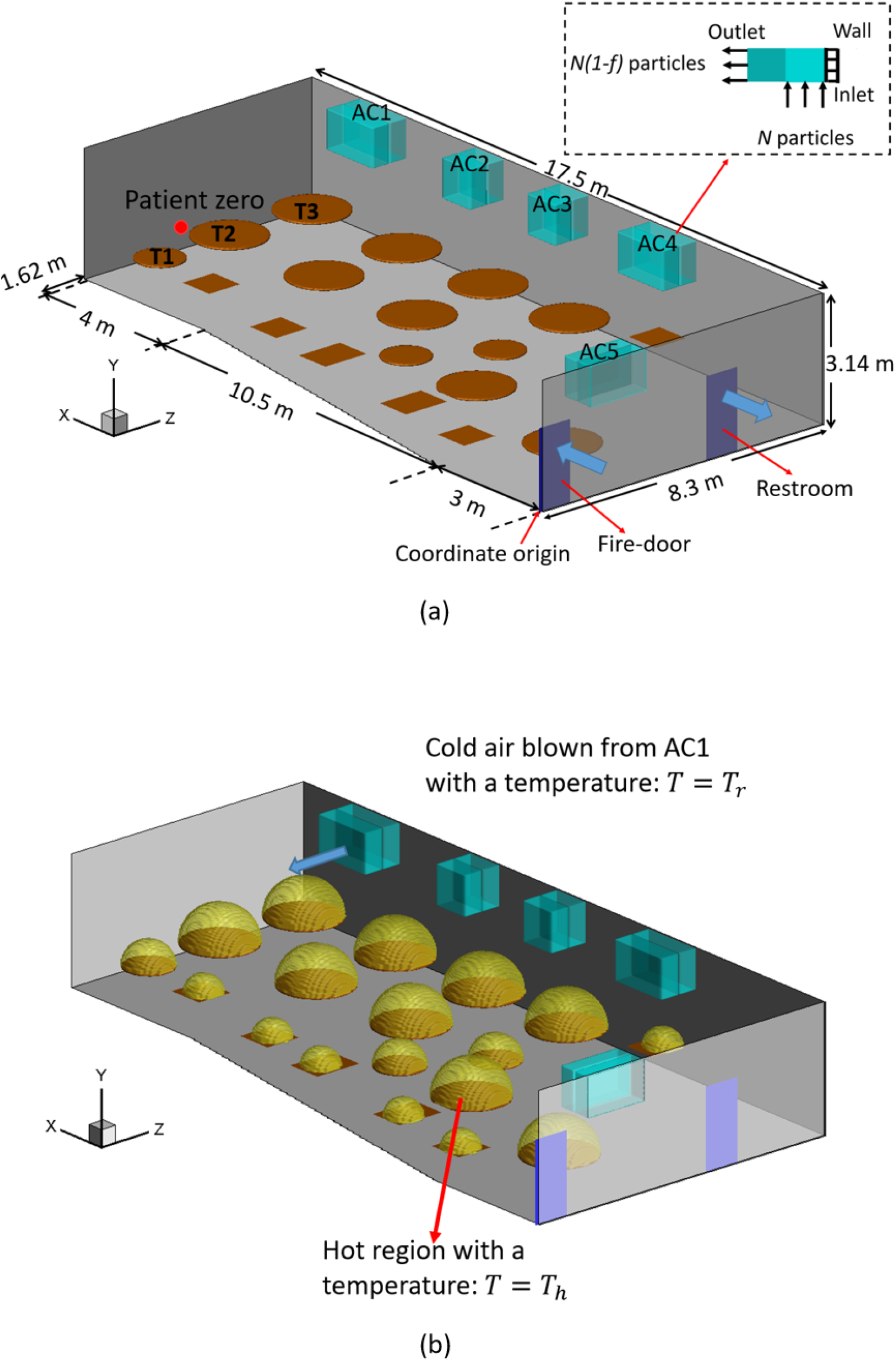
Sketch of the simulation set-up: (a) settings of air conditioners (AC), doors, and tables, and (b) modelling of thermal effect. The grey shades show the walls. The cyan rectangular boxes show the air conditioners, which are labeled as AC1, AC2, AC3, AC4, and AC5 in (a). The brown thin boxes and cylinders show the rectangular and round table surfaces. The dark blue rectangles on the wall show the fire-door and the door to a restroom. The tables with infections are labeled as T1, T2, and T3 in (a). The zoom-in region in (a) shows the inlet, outlet, and filtration setting of the ACs. The blue arrows close to the fire-door and the restroom in (a) show the flow directions at the two doors, respectively. The yellow hemispheres in (b) show the hot region.

**FIG. 2.**
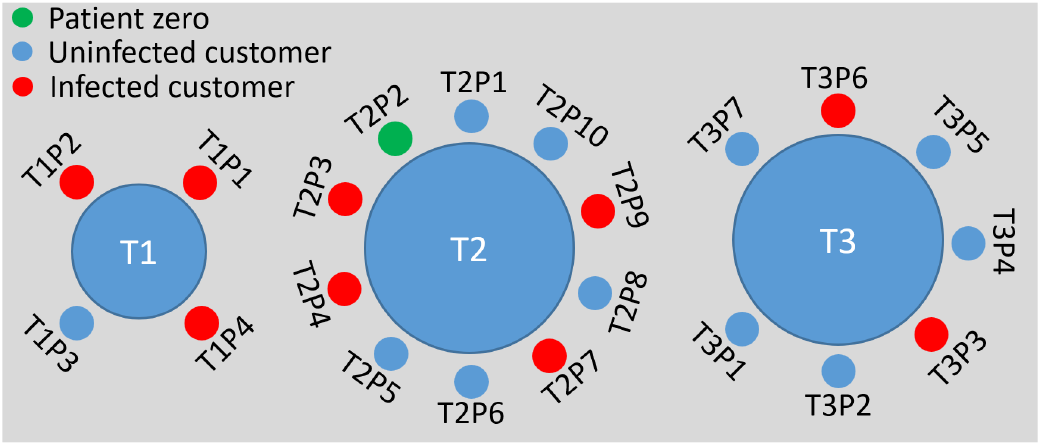
Sketch of the customers sitting around table T1, T2, and T3 during the lunch time of January 24^*th*^, 2020, in the Guangzhou restaurant shown in Fig. 1.

Our numerical simulations are set up with the configuration shown in Fig. 1. A three-dimensional Cartesian coordinate system is used in the mathematical formulation and numerical discretization of the computational domain, in which the *x*-direction is set parallel to the longer wall of the room, the *y*-direction is set as the opposite of the direction of gravity, and the *z*-direction is set parallel to the shorter wall of the room. The origin is located at a corner shown in Fig. 1(a). The length of the longer wall is 17.5 m, which is located at *z* = 8.3 m. The lengthes of the two shorter walls are 8.3 m and 6.68 m, which are located at *x* = 0 m and *x* = 17.5 m, respectively. The other wall has three segments as shown in Fig. 1(a). The tables in this restaurant are captured in the simulation using the IB method. There are two types of table, one is round-shaped and the other is rectangular-shaped. The round-shaped tables have two sizes with diameters *d* = 1.2 m and *d* = 1.8 m, respectively, while the rectangular-shaped tables have two sizes as 0.9 m *×* 0.9 m and 1.2 m *×* 0.9 m. The height of all the tables in the simulations is set as *h*_*t*_ = 0.75 *m*, based on our understanding of the typical height of the tables used in China.

There are five working HVAC (heating, ventilation, and air conditioning) systems in this room as shown in Fig. 1(a), which are labeled in short as AC1, AC2, AC3, AC4, and AC5, and are also simulated by IB method. Four of them are installed on the wall at *z* = 8.3 m, and the other one is installed on the wall at *x* = 0 m. AC1, AC4, and AC5 have a size of 1.6 m *×*0.5 m *×*0.25 m (length *×* width *×* height), while AC2 and AC3 have a size of 1 m *×* 0.5 m *×* 0.25 m. Each AC has both an inlet and an outlet, as illustrated in Fig. 1(a). At the inlet, air flow is sucked into the AC in the *y*−direction (i.e. upward vertically) with a ventilation rate *q*_AC_. At the outlet, air flow is blown out of the AC horizontally with the same ventilation rate *q*_AC_. On the wall at *x* = 0 m, two rectangular regions [colored by dark blue in Fig. 1(a)] are used to simulate the fire-door and the exhaust vent in the restroom of this restaurant, respectively. Fresh air is sucked into the restaurant room through the fire-door, while the polluted air exits through the restroom. The set-up of the restroom is simplified since virus-laden droplets are mostly concentrated near AC1. The ventilation rates of the fire-door and the restroom are both set as *q* = 0.044 m^3^*/*s following the literature^9^.

To simulate the thermal effect by the human bodies around the tables and the hot food on the tables, a simplified model is implemented, as illustrated in Fig. 1(b). Above each table, a hemispherical hot region is defined, which has a diameter the same as that of the round table below it, or the diagonal length of the rectangular table below it. In each hot region, the temperature is set by a constant *T* = *T*_*h*_. Moreover, the temperature of the air blown out of the AC outlets is set to be a constant *T* = *T*_*r*_, which is lower than *T* = *T*_*h*_. The temperature difference between them is denoted by Δ*T* = *T*_*h*_ − *T*_*r*_. The dimensionless temperature

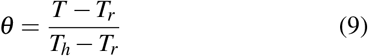

is used to quantify the temperature variation in the flow field. Adiabatic condition is applied at the walls, floor, and ceiling. The initial temperature in the room is set as *T* = *T*_*r*_ in the simulation.

In the simulation, the height of the mouths of all the customers is set as *h*_*m*_ = 1.3 m, based on the statistics of the averaged height of the sitting people^38^. Virus-laden aerosols are released from patient zero, which is marked by the red dot in Fig. 1(a). aerosols are initialized following the measurement by Shao *et al*.^39^. It is assumed that each breath cycle of patient zero includes a 2.37 s inhale period and a 1.58 s exhale period. During the exhale period, the aerosols are released uniformly in a circular disk with a diameter of 40 mm, which is a size based on the averaged diameter of mouths^40^. The velocity of the aerosols is set as the same as the breathing flow, which is pointing to the center of T2. In the simulations, there are 44 aerosols released during each exhale period, based on the measurement result^39^. The diameter of the aerosols is set as 1.5 *µ*m, following the peak value of the measured size distribution^39^. The density of the aerosols is set to be the density of water as 10^3^ kg*/*m^3^, since the virus-laden aerosol being simulated is a droplet. Based on the research by Alsved *et al*.^41^, the number of the aerosols released per second by normal talking and that by loud talking are respectively about two and four times as that by normal breathing. In order to consider the effect by people normal talking or loud talking in the restaurant, cases with higher numbers of aerosols released per breath cycle have also been simulated, which produced similar qualitative results compared with the cases using the normal breathing setting. In the following sections, only the results from the normal breathing setting are reported.

The filtration effect of the ACs and aerosol deposition are considered in our simulations. As shown in Fig. 1(a), the filtration effect is quantified by the filtration efficiency *f*. Once *N* aerosols have been sucked into an AC, the *N*(1 − *f*) of them will be re-ejected into the room randomly from the outlet of the AC. In the simulation, the filtration efficiency is set as *f* = 80%, based on the typical performance of HVAC systems^42^. To simulate the deposition effect, an aerosol will be labeled as deposited and deleted from the computational domain once the distance between the aerosol and the solid surface close to it (walls, tables, and ACs) is smaller than the radius of the aerosol.

### C. Simulated cases

The computational domain in the present study has a grid resolution of *N*_*x*_ *× N*_*y*_ *× N*_*z*_ = 200 *×* 200 *×* 200. The Reynolds number is defined as

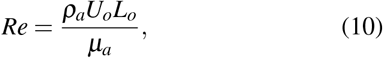

and has the value *Re* = 6.8 *×* 10^4^, where *ρ*_*a*_ is the density of air, *U*_*o*_ = 1 m*/*s, and *L*_*o*_ = 1 m*/*s, Four cases with various ventilation rate *q*_AC_ and temperature difference Δ*T* are considered as shown in Table I. The range of *q*_AC_ covers the common range of the ventilation rate of a HVAC system^43^. The temperature difference is based on the consideration of the human body temperature, the food temperature, the room temperature, and the implementation of the simplified model shown in Fig. 1(b).

**TABLE I.** Simulation cases and parameters.

| Cases | $q_{AC} \text{ (m}^3/\text{s)}$ | $\Delta T \text{ (}^\circ\text{C)}$ |
| --- | --- | --- |
| AC14DT00 | 0.14 | 0.0 |
| AC14DT10 | 0.14 | 10.0 |
| AC112DT00 | 1.12 | 0.0 |
| AC112DT10 | 1.12 | 10.0 |

The simulations were run with sufficient duration for the flow field to fully develop. Figure 3 shows the time evolution of the velocity variances ⟨*u*′ *u*′⟩, ⟨*v* ′*v*′⟩, and ⟨*w* ′*w*′⟩ in case AC112DT10, where *u*′, *v*′, and *w*′ are defined in Eq. (13), and ⟨*·*⟩ is the spatial–temporal average defined as

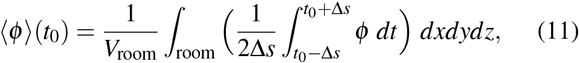

where *t*_0_ − Δ*s < t < t*_0_ + Δ*s* is a time window expanding from *t* = *t*_0_ that is long enough to cover the temporal fluctuations, *V*_room_ is the whole volume of the room. The aerosols are released after the flow in the room has fully developed, as illustrated in Fig. 3. Then the simulations continued till the total number of aerosols in the whole computational domain is statistically converged as a result of the balance between the number of newly released aerosols and that of deposited, filtered and exiled aerosols.

**FIG. 3.**
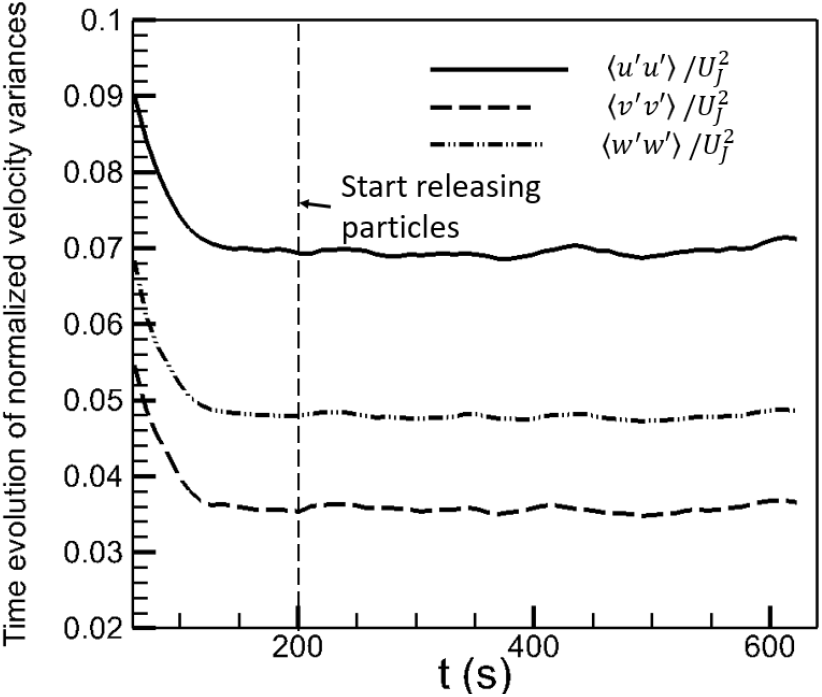
Time evolution of velocity variances ⟨*u′ u′*⟩, ⟨*v′ v′*⟩, and ⟨*w′ w′*⟩, normalized by the jet velocity *U*_*J*_ at the outlet of AC1, in case AC112DT10.

## III. RESULTS AND DISCUSSIONS

### A. Flow structures under various indoor conditions

In this subsection, the indoor airflow structures are discussed to provide a general picture of the flow field in the simulated cases. The distributions of mean flow structures and turbulent flow structures under different ventilation rates and thermal settings are illustrated, which will be connected to the aerosol exposure index and the infection risk of COVID19 discussed in Secs. III B and III C, respectively.

To investigate the indoor flow structure, a temporal average is defined as

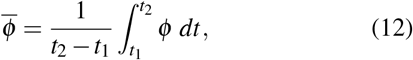

where *t*_1_ is a selected time instance after the total number of aerosols reaches dynamical balance in the simulation. Based on the temporal average, the flow field is decomposed into two parts: the mean flow and the turbulence,

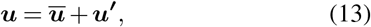

where ***ū*** is the mean flow velocity and ***u***′ is the turbulent velocity fluctuation.

Figure 4 shows the streamlines of the mean flow traced forward from the outlets of the ACs, under different ventilation rates and thermal settings. The flow structures in the cases without the consideration of thermal effect are displayed in Figs. 4(a) and (c). It is observed that a recirculation flow with the size of the width of the room is formed in front of the ACs in cases AC14DT00 and AC112DT00, where the air ejected from the ACs first moves along the initial jet direction, turns to the floor direction once it reaches the wall, moves downward until reaching the floor, turns back towards the ACs, moves under the tables, and in the end, is sucked into the ACs through the inlets. In the cases with the consideration of thermal effect, the flow structures are more complex, as shown in Figs. 4(b) and (d). It is found that the cold air blown from the ACs moves downward at an angle due to the interaction between the cold jet of a higher density and the rising thermal plume of a lower density near T3. Before reaching the floor, the cold air is heated by the interaction. Comparing cases AC14DT10 and AC112DT10, it is illustrated in Fig. 4(b) that in case AC14DT10, the heated air near the floor that originates from the jet from AC1 moves in various directions. In case AC112DT10 [Fig. 4(d)] with a stronger jet momentum, the air from AC1 moves in the initial jet direction till reaching the wall near *z* = 0 m. For the same reason, the jet inclination angle in case AC14DT10 is larger than that in case AC112DT10.

**FIG. 4.**
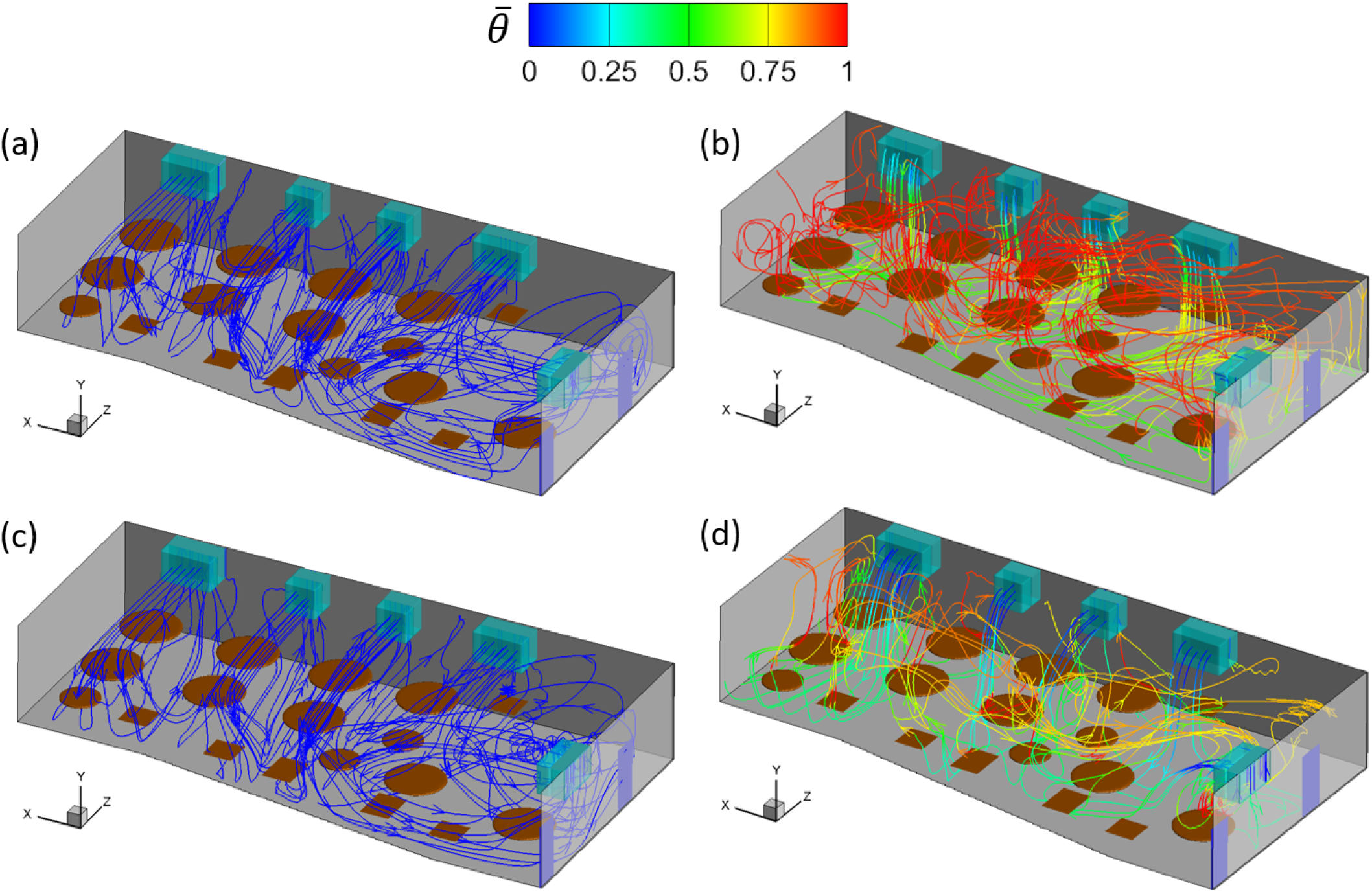
Mean flow streamlines traced forward from the outlets of the ACs in the simulation cases: (a) AC14DT00, (b) AC14DT10, (c) AC112DT00, and (d) AC112DT10. The streamlines are colored by the local mean temperature 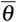.

The exchange between the indoor and outdoor airs is investigated. To reduce the concentration of virus-laden aerosols, besides applying the filtration system, the most commonly used method is to keep exchanging indoor and outdoor airs. Fresh outdoor air can reduce the indoor aerosol concentration through the dilution effect. Figure 5 shows the streamlines of the mean flows traced backward from the restroom. As shown, the range covered by these streamlines varies with indoor conditions. The shortest range is from the tables in front of AC4 to the restroom, which occurs in cases AC14DT00 and AC112DT00, while the longest range is from the space in front of AC1 to the restroom, which happens in case AC112DT10. Figure 5 also shows that there are barely any streamlines near T1-3 that are directly connected to the restroom and thus it is less likely for the aerosols in this area to exit the restaurant, which is consistent with the measurement by Li *et al*.^9^.

**FIG. 5.**
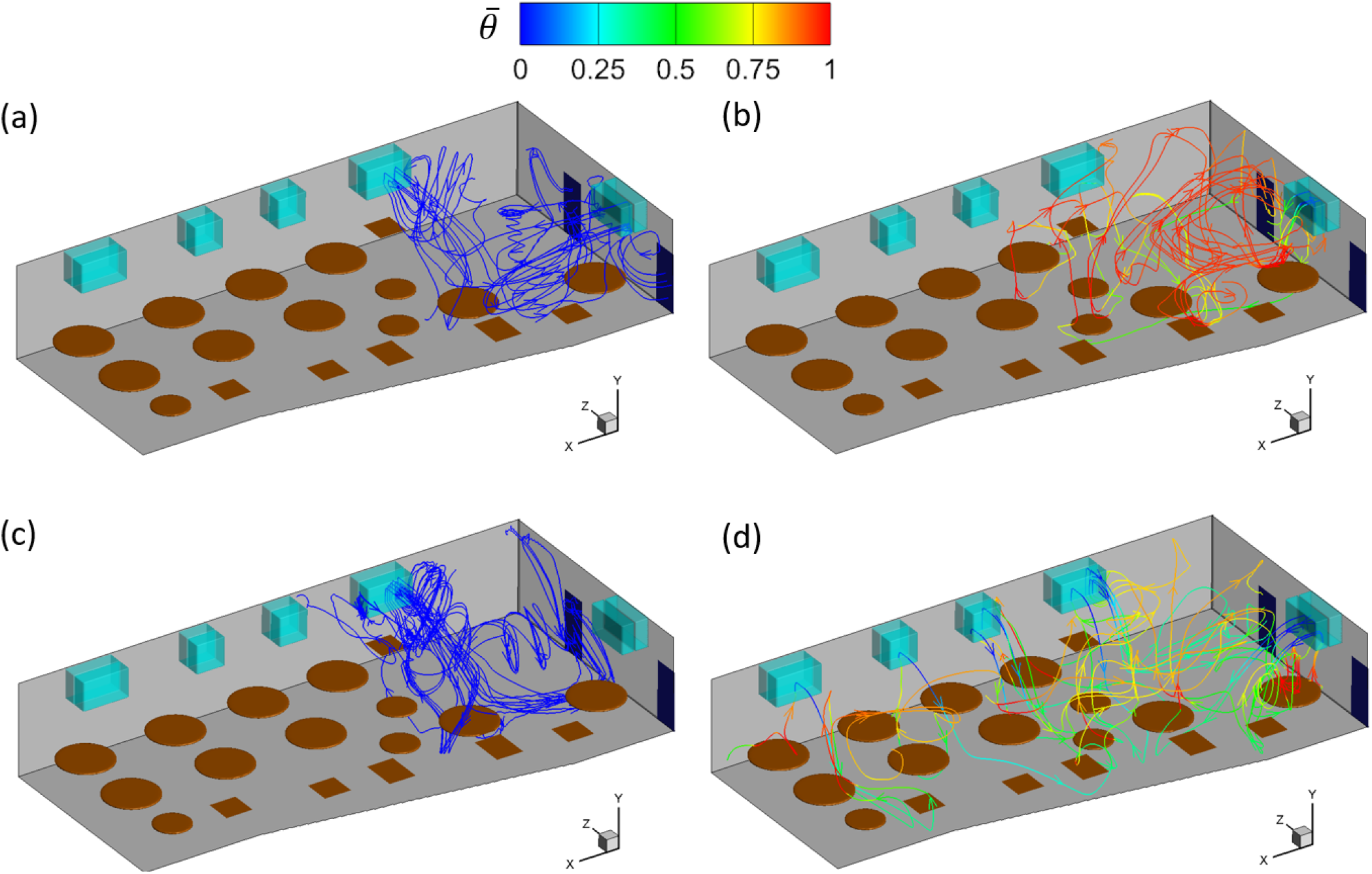
Mean flow streamlines traced backward from the restroom in the simulation cases: (a) AC14DT00, (b) AC14DT10, (c) AC112DT00, and (d) AC112DT10. The streamlines are colored by the local mean temperature 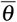.

The turbulence is quantified by the turbulent kinetic energy (TKE), defined as

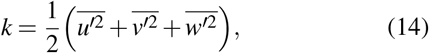

and is displayed by the contours plotted near T1-3 in Fig. 6. In Figs. 6(a) and (c), it is illustrated that the turbulence is mostly concentrated around the jet from AC1, in the two cases without considering thermal effect. Moreover, the TKE in Fig. 6(c) is higher than that in Fig. 6(a) in most of the displayed area, which is caused by the stronger entrainment by the jet in case AC112DT00 than in case AC14DT00. However, the distribution of turbulence is different in the cases with the consideration of thermal effect. In Figs. 6(b) and (d), it is found that the turbulence is mostly concentrated near the region where cold air interacts with the thermal plume, which is located above T3 in case AC14DT10 and has a broader range in case AC112DT10. Relatively weaker turbulence is also observed from Figs. 6(b) and (d) above the surfaces of T1 and T2, which is produced by the instability of the rising thermal plume. Comparing Figs. 6(b) and (d), it is shown that the turbulence in case AC112DT10 is stronger than that in case AC14DT10 in most of the area, due to the stronger interaction between the cold jet and the thermal plume in case AC112DT10. Furthermore, comparing Figs. 6(b) and (d) with Figs. 6(a) and (c), it is found that the cases with the consideration of thermal effect show more intense turbulence near T1-3 than the cases without the thermal effect. In summary, different ventilation rates and thermal settings give rise to different distributions and intensity levels of the indoor turbulence. Another conclusion that is not directly shown from these results but is reasonable to expect is that the relative location between the tables and ACs also impacts the distribution of indoor turbulence.

**FIG. 6.**
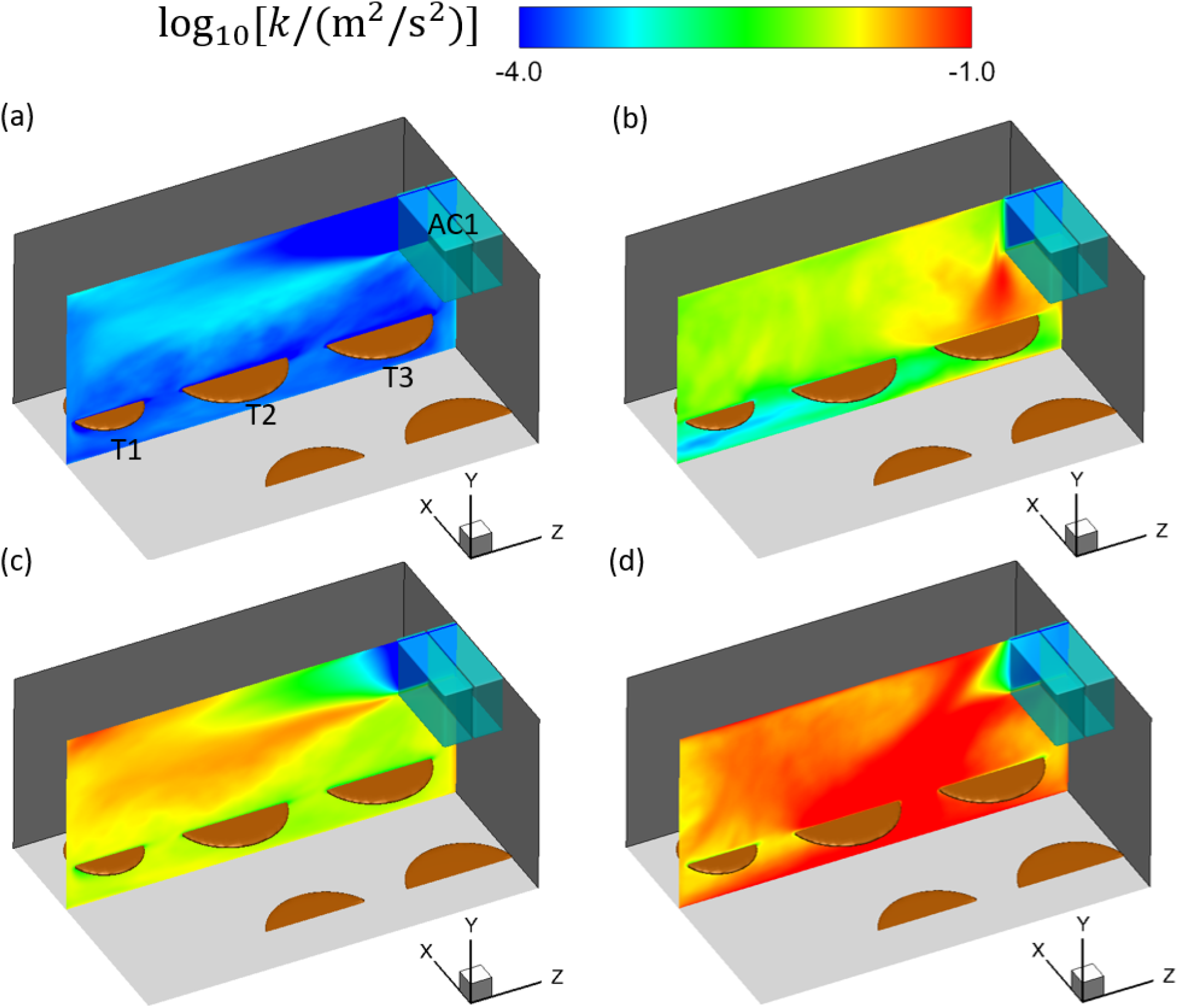
Contours of log_10_[*k/*(m^2^*/*s^2^)] in the plane at *x* = 13 m in the simulation cases: (a) AC14DT00, (b) AC14DT10, (c) AC112DT00, and (d) AC112DT10.

### B. Exposure index of virus-laden aerosols

Before using the simulated data to predict the COVID-19 infection risk through airborne transmission, first, in this section, we illustrate and analyze the distributions of aerosols under different ventilation rates and thermal settings. Explanation on the aerosol distribution is also given in this section, based on the flow structure illustrated in Sec. III A.

The indoor spatial–temporal distribution of the aerosols under various ventilation rates and thermal settings is quantified by the aerosol concentration *C*(***x***, *t*), which is defined as the number of aerosols per unit volume. The mean value of *C* during the statistical steady state, which is also called aerosol exposure index in this study, is used to quantify the temporally averaged distribution of aerosols or the degree of being exposed to the aerosols in the restaurant, which is numerically calculated in the present simulations as

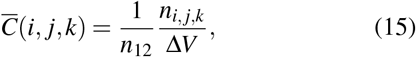

where *n*_*i, j,k*_ is the total number of aerosols that have been in the grid cell (*i, j, k*) at each timestep during the time period *t*_1_ *< t < t*_2_, Δ*V* is the volume of the grid cell, and *n*_12_ is the total time steps for the simulations running from *t*_1_ to *t*_2_.

In the simulations, the aerosols are transported by three forces on the right hand side of Eq. (6), i.e. the Maxey–Riley equation, including the Stokes drag force, the gravitational force, and the Brownian motion effect force. It is found from the results that in the simulated cases, the Stokes drag force ***f***_stokes_ is dominant over the other two forces in most of the computational domain. It can be further decomposed into four parts by substituting Eq. (13) into Eq. (6),

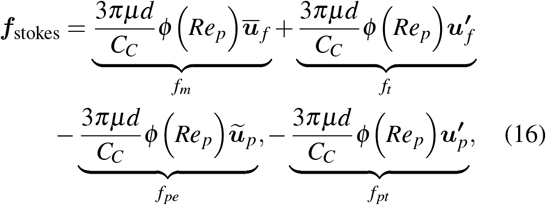

where *ũ* is the ensemble averaged velocity of the aerosol, ***f***_*m*_ is the force due to the mean flow velocity, ***f***_*t*_ is the force due to the turbulent fluctuation, ***f***_*pe*_ is the force due to the mean velocity of the aerosol, and ***f***_*pt*_ is the force due to the fluctuating velocity of the aerosol. Based on the above decomposition, an aerosol in general experiences less Stokes drag if its mean velocity *ũ*_*p*_ is closer to the local mean flow velocity *ū*_*f*_, because of the opposite effects of ***f***_*pe*_ and ***f***_*m*_. Therefore, the mean flow tends to drag an aerosol to move along with its streamline by the combined effect of ***f***_*m*_ and ***f***_*pe*_. The local turbulence tends to randomly disperse a group of aerosols^44–48^ through the fluctuating shear forces ***f***_*t*_ + ***f***_*pt*_. The aerosol exposure index in the statistically steady state results from the combination of all these effects.

Figure 7 shows the iso-surfaces of aerosol exposure index 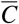 for 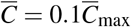 (in black) and 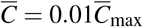 (in yellow) under different ventilation rates and thermal settings, where 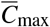 is the maximum of 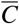. Comparing the iso-surfaces of 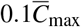 and 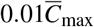, we can see that the former is closer to patient zero and located around T2, while the latter reaches T1 and T3 in cases AC14DT00, AC112DT00, and AC14DT10. In case AC112DT10, it is observed from Fig. 7(d) that the iso-surface of 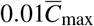 is preferentially concentrated around T3, in contrast to T1. The values of *C*_max_ in the different cases are: *C*_max_ = 2740.9 in case AC14DT00, *C*_max_ = 783.6 in case AC14DT10, *C*_max_ = 276.9 in case AC112DT00, and *C*_max_ = 279.3 in case AC112DT10. Therefore, there are more aerosols concentrated inside the iso-surface of 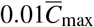 in case AC14DT00 than the other three cases, because the stronger air flows due to the higher *q*_AC_ and thermal effect in cases AC14DT10, AC112DT00, and AC112DT10 drive more aerosols to collide and deposit on the walls near T1-3, so that the total numbers of aerosols have lower values in the latter three cases. Comparing the range reached by the iso-surfaces of 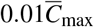 in the *x*−direction with that reached in the *z*−direction, we can observe that in all the cases the aerosol exposure index decays faster along the *x* − direction than in the *z* − direction, since the mean flow drag *f*_*m*_ is dominant in the *z*−direction over in the *x*−direction due to the jet flow from AC1. Moreover, the topography of the iso-surface of 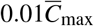 varies with the ventilation rates and thermal settings. Figures 7(a) and (c) display an L-shaped contours of 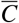 inside the iso-surface of 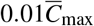 in cases AC14DT00 and AC112DT00. The lower aerosol exposure index in the region between the two sides of the L-shape is due to the turbulent dispersion around the jet flow. Figures. 7(b) and (d) show a more complex distribution of the aerosol exposure index with a rougher topography of the iso-surface of 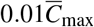 in cases AC14DT10 and AC112DT10, because the rising plumes due to the thermal effect shown in Figs. 4(b) and (d) create a more complex drag force field. Moreover, Fig. 7(d) shows a preferential concentration on the T3 side than on the T1 side due to the transport by the stronger mean flow in case AC112DT10 compared with case AC14DT10.

**FIG. 7.**
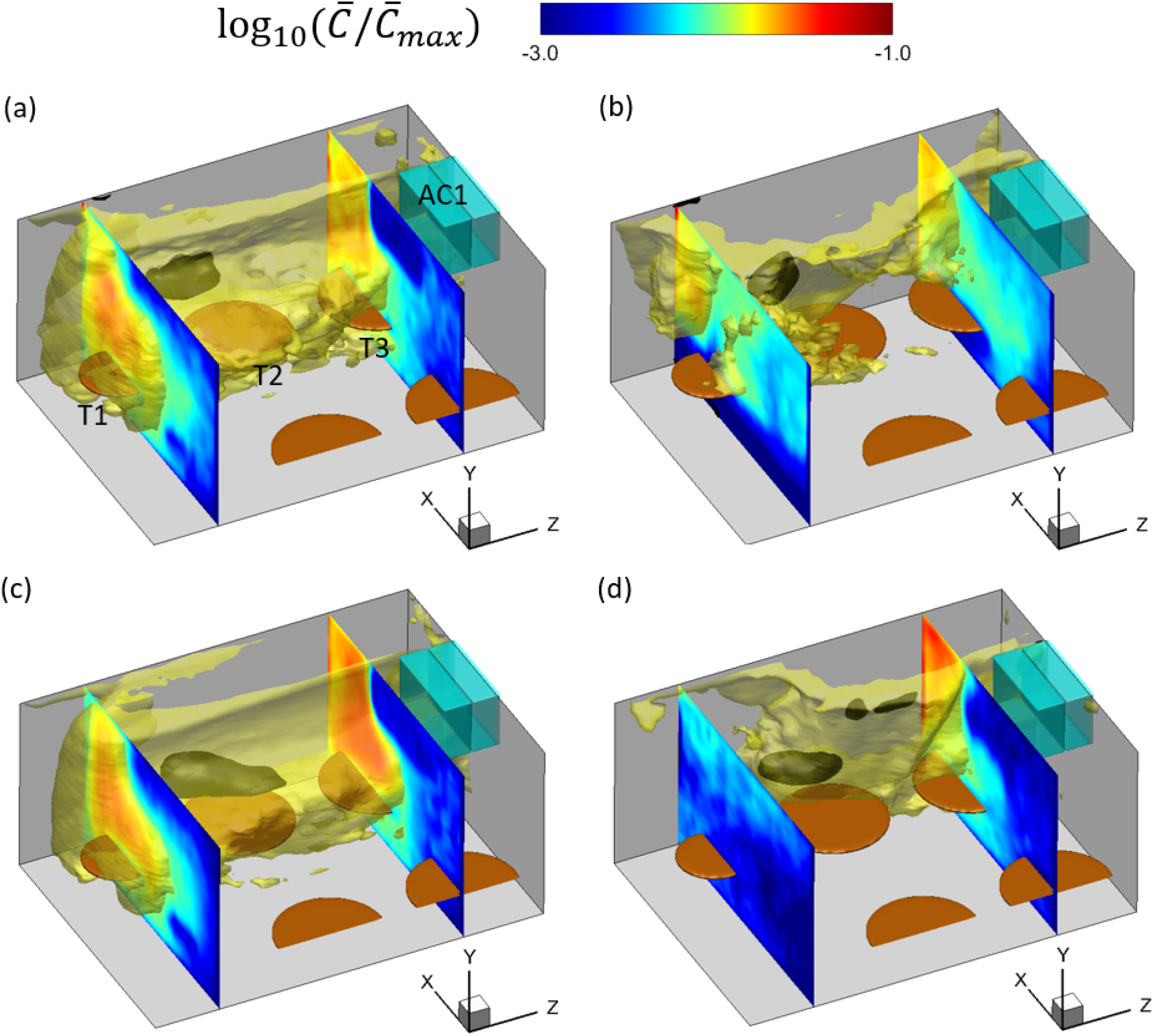
Iso-surfaces of 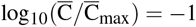 (black) and 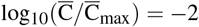 (yellow), and the contours of 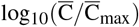 in the planes at *z* = 2.6 m (across T1) and *z* = 6.8 m (across T3), in the simulation cases: (a) AC14DT00, (b) AC14DT10, (c) AC112DT00, and (d) AC112DT10. Here, 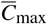 is the maximum of 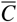.

Since the infections at T1 and T3 are believed to be directly related with the airborne transmission^9^, more research attentions are paid to the regions close to T1 and T3. The iso-surface of 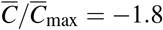 (the threshold is selected as a representative one based on the isolines concentrated near T1 and T3, see Figs. 9 and 11) and the mean flow streamlines near T1 and T3 are shown in Fig. 8 and Fig. 10, respectively. The isolines of 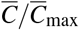 near T1 and T3 at the height of mouth *h*_*m*_ are also displayed in Fig. 9 and Fig. 11, respectively. The correlations between the contours of aerosol exposure index and the mean flow streamlines are evident.

**FIG. 8.**
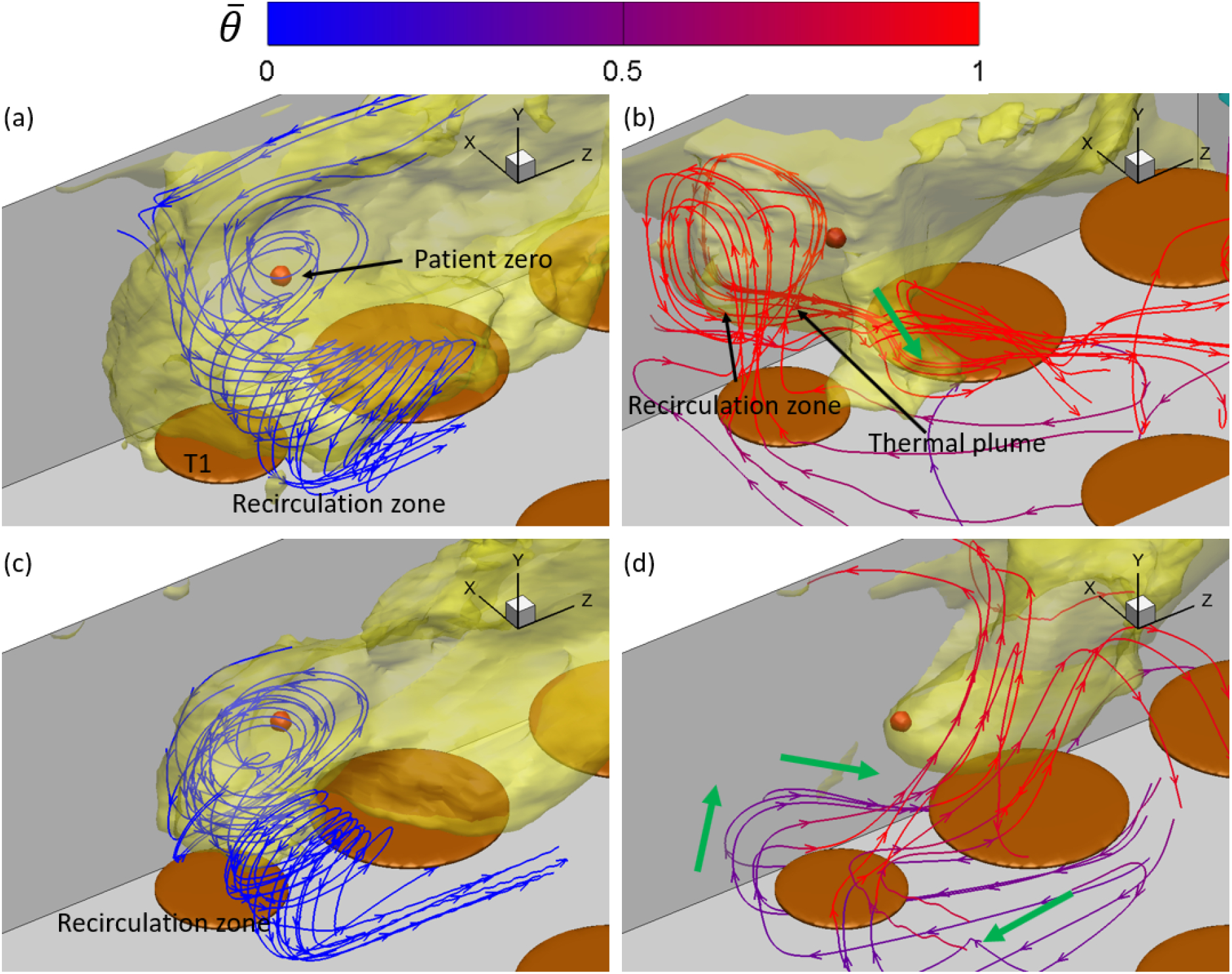
Iso-surface of 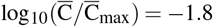 (yellow) around T1 and the mean flow streamlines colored by the local mean temperature 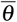, in the simulation cases: (a) AC14DT00, (b) AC14DT10, (c) AC112DT00, and (d) AC112DT10. Here, 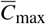 is the maximum of 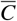.

**FIG. 9.**
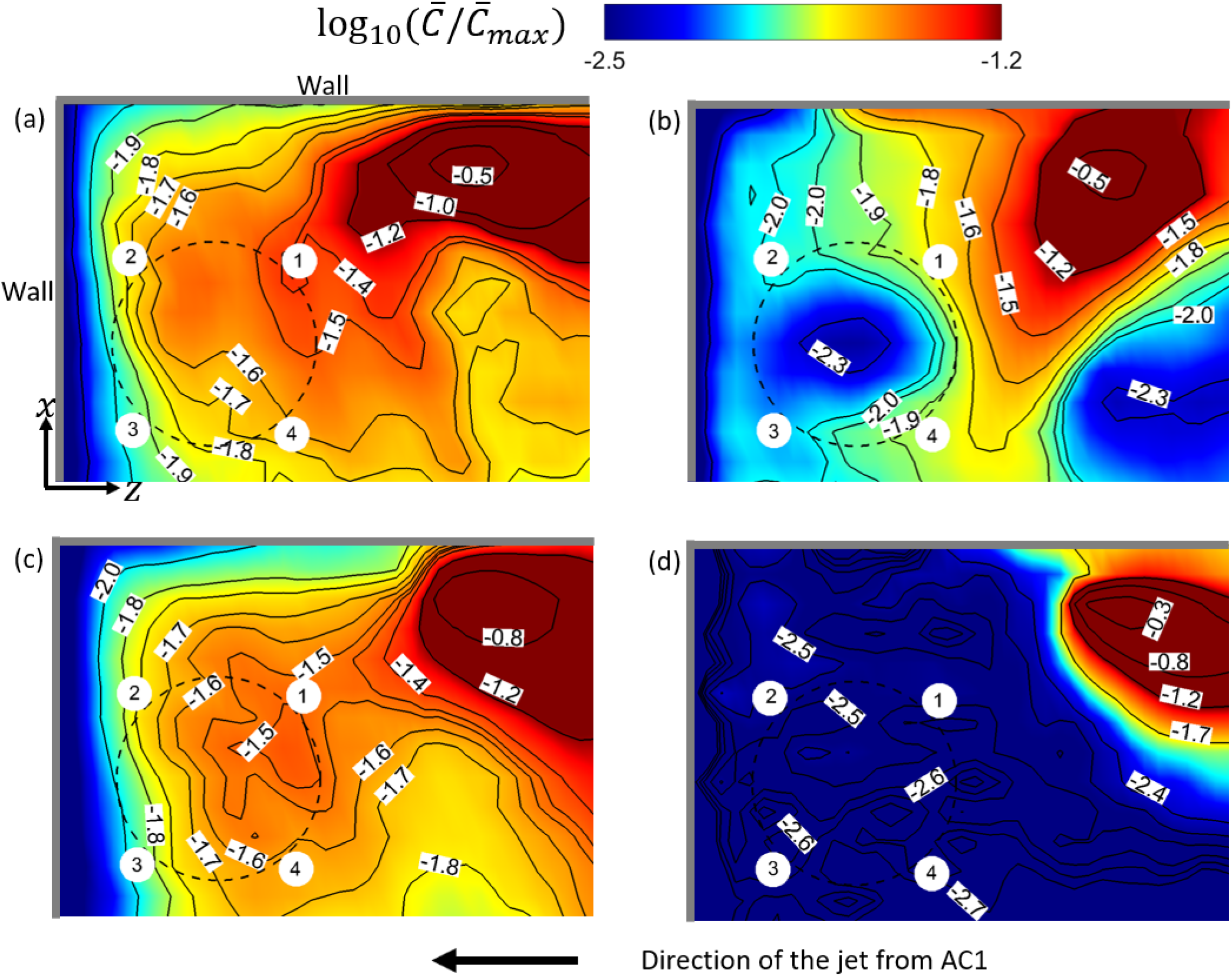
Contours of 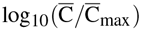 at *y* = 1.3 m and the corresponding isolines near T1, in the simulation cases: (a) AC14DT00, (b) AC14DT10, (c) AC112DT00, and (d) AC112DT10. T1 is denoted by the black dashed circle, and the people sitting around it are displayed by the small white circular regions.

**FIG. 10.**
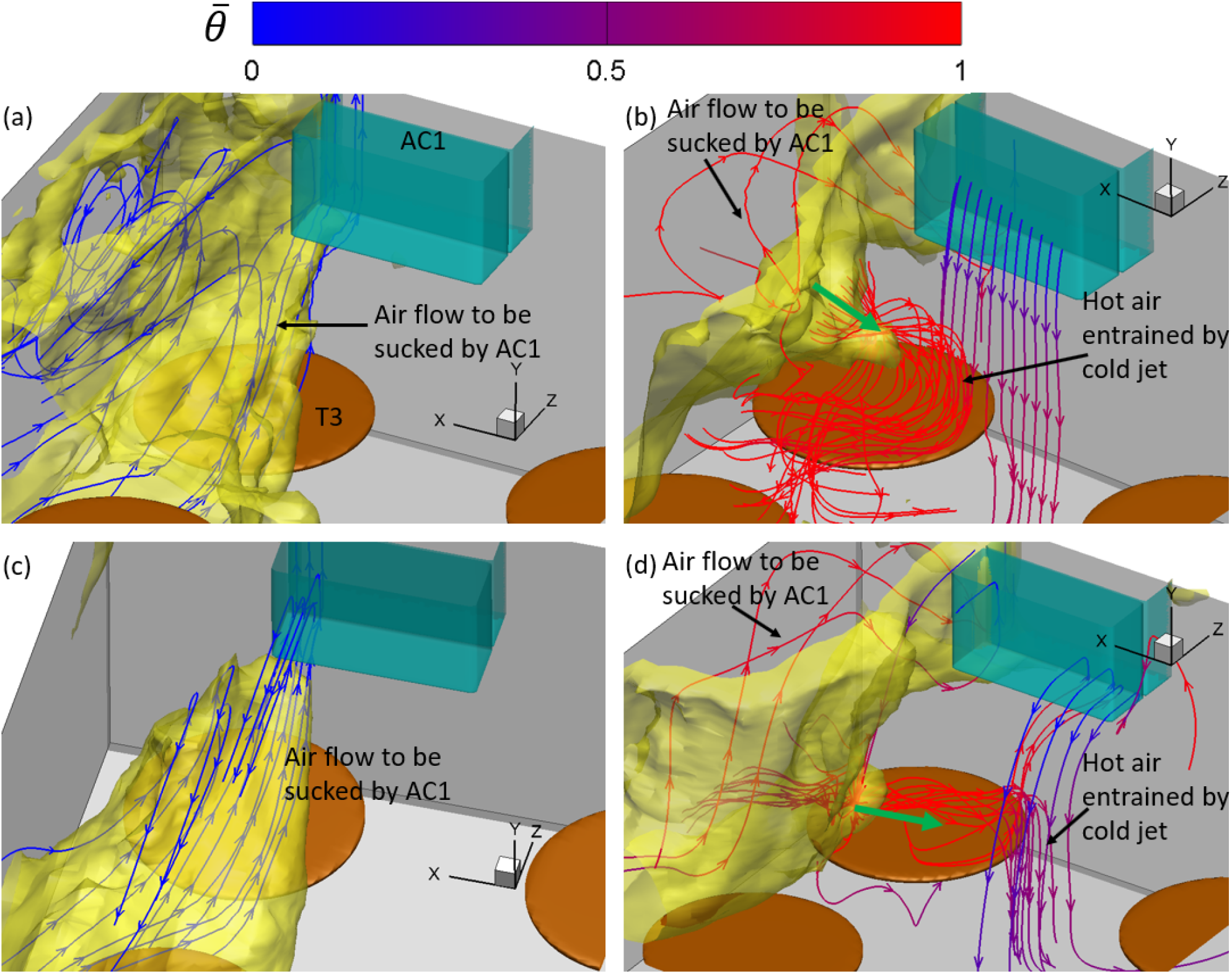
Iso-surface of 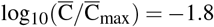 (yellow) around T3, where 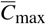 is the maximum of 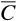, and mean flow streamlines colored by the mean temperature 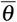, in the simulation cases: (a) AC14DT00, (b) AC14DT10, (c) AC112DT00, and (d) AC112DT10.

**FIG. 11.**
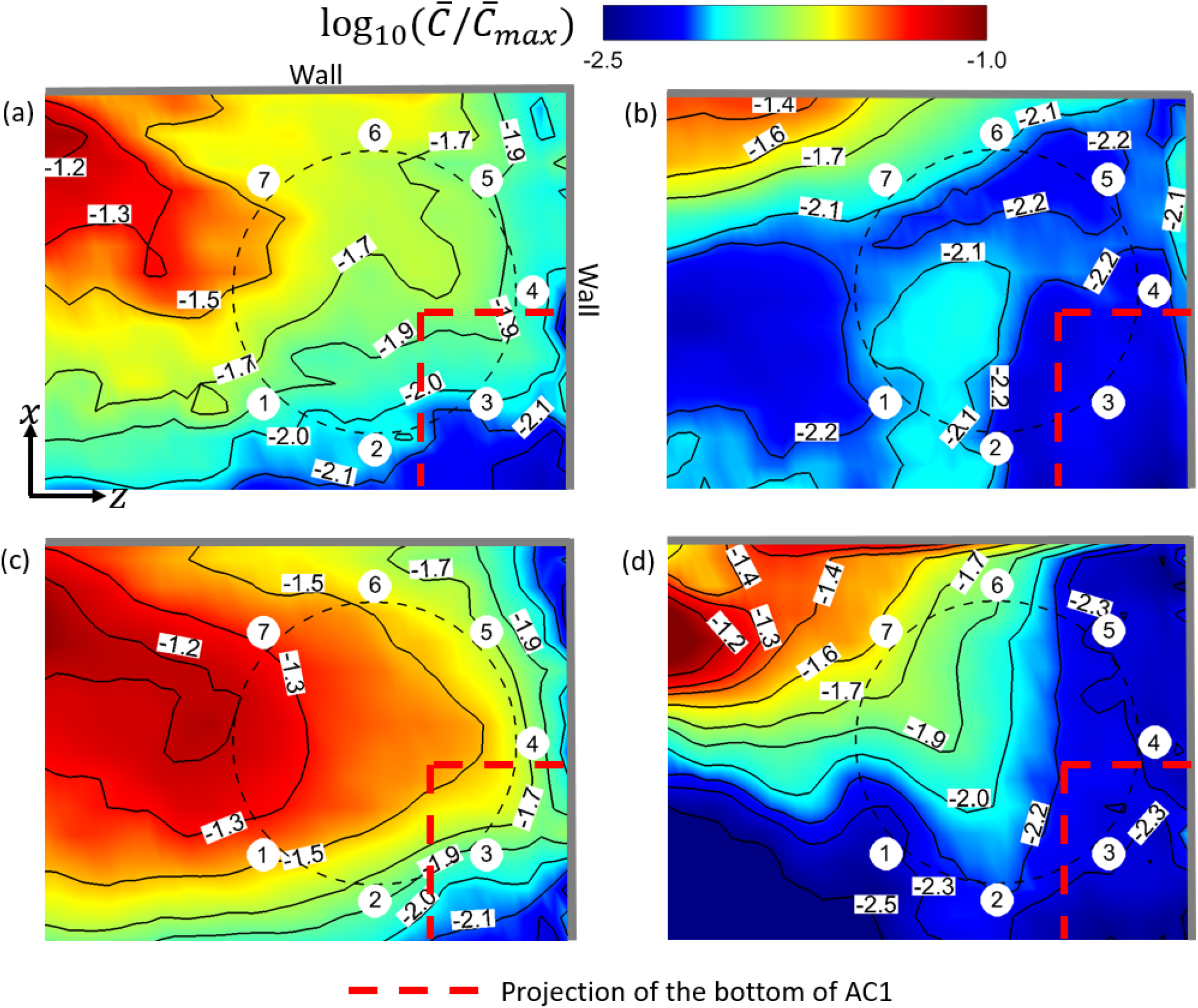
Contours of 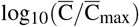 at *y* = 1.3 m and the corresponding isolines near T3, in the simulation cases: (a) AC14DT00, (b) AC14DT10, (c) AC112DT00, and (d) AC112DT10. T3 is denoted by the black dashed circle, and the people sitting around it are displayed by the small white circular regions.

Near T1, the mean flow streamlines in Figs. 8(a) and (c) indicate that there exists a recirculation zone above T1 in cases AC14DT00 and AC112DT00. Inside the recirculation zone, the streamlines originate from the region around T2, first move towards the negative *z*−direction, and then gradually turn to T1 with a spiral-like trajectory. The recirculation zone above T1 is generated by the interaction among the walls, the surface of T1, and the local air flows. It is illustrated in Figs. 9(a) and (c) that from 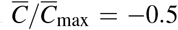 to −1.8, the isolines of 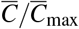 near T1 tend to be elongated while turning to T1. This pattern of spreading is due to the convection by the mean flow in the recirculation zone observed in Figs. 8(a) and (c). In Fig. 8(b), two peaks on the iso-surface of 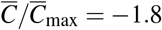 are observed around T1 in case AC14DT10. The left peak is associated with a recirculation zone attached to the wall, which is formed by the thermal plume rising from T1 indicated by the red streamlines. Another peak is located between T1 and T2, where hot air passes through, indicated by the streamlines together with the green arrows in the Fig. 8(b). Corresponding to the two peaks observed in Fig. 8(b), it is illustrated in Fig. 9(b) that from 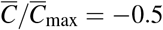 to −2, the isolines of 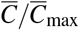 are preferentially distributed in the region between T1 and the wall, where there is the recirculation zone, and in the region between T1 and T2, where the hot air flow caused by the thermal passes by. The spreading of aerosols to the first region is due to the convection by the thermal plume that generates the recirculation, and the spreading to the latter region is due to the convection by the hot air flow passing by. As for the case AC112DT10, it is found in Fig. 9(d) that the magnitude of the gradients of aerosol exposure index 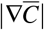 around the isoline of 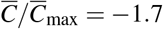 is significantly larger than those in the other cases, which indicates the less spreading of the aerosols in the region colored by dark blue in Fig. 9(d). The less spreading is consistent with the observation in Fig. 8(d). Near T1, the streamlines are directly connected to AC1, indicating that the aerosols in the dark blue region in Fig. 9(d) are mostly transported by the mean flow along these streamlines. However, there are less aerosols concentrated in front of AC1 as shown in Fig. 7(d). Therefore, less aerosols are transported to the region near T1 in case AC112DT10. The irregular distribution of the isolines of 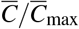 in Fig. 9(d) also indicates that the turbulence due to the thermal effect around T1, which is also illustrated in Fig. 6(d), plays a role in dispersing the aerosols by exerting the turbulent shear force ***f***_*t*_ + ***f***_*pt*_ on the aerosols.

To illustrate the aerosol exposure index near T3, Figs. 10(a) and (c) show the streamlines of the mean flow inside the iso-surface of 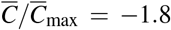 in cases AC14DT00 and AC112DT00. Along these streamlines, the air moves from the region close to patient zero to the wall and the inlet of AC1. The isolines of 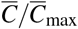 displayed in Figs. 11(a) and (c) are elongated in the same direction around T3, from 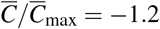 to 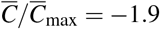. The elongation indicates the aerosol spreading, which is due to the transport by the mean flow shown in Figs. 11(a) and (c). Comparing Fig. 11(a) with (c), the isolines in the latter figure are sharper near the spot below AC1. This is because the stronger suction in case AC112DT00 exerts more forcing on the aerosols, which attracts more aerosols to AC1 compared with case AC14DT00. As shown in Figs. 11(b) and (d), in both the cases AC14DT10 and AC112DT10, there exists a peak of the iso-surface of 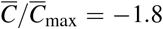, which is located above the surface of T3. The isoline of 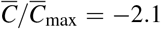 in Fig. 11(b) indicates a local high aerosol exposure index region, which corresponds to the peak observed in Fig. 10(b) in case AC14DT10. Also in Fig. 11(d), from 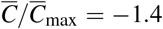 to 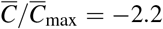, the elongation of the isolines above T2 corresponds to the peak observed in Fig. 10(d). The streamlines in parallel with the green arrows in Figs. 10(b) and (d) indicate that the aerosol spreading in this region is due to the mean flow convection associated with the heated air to interact with the cold air in the jet above T3. Comparing the peak observed in case AC14DT10 with the one observed in case AC112DT10, the latter is located lower, because in the latter case, the cold jet from AC1 has stronger momentum, which makes the converging point closer to the surface of T3.

### C. Prediction of infection risk based on the simulated results

In this section, first, a discussion about using the simulated results to predict the infection risk of COVID-19 is presented. Then, the simulation-based distribution of the infection risk is illustrated and explained by linking it to the analysis in Sec. III B.

There are various factors that can determine the total infection risk of COVID-19 associated with airborne transmission. According to Mittal *et al*.^50^, the total infection risk *R*_in_ is the product of the risk functions of a series of complex factors and processes, which can be expressed as

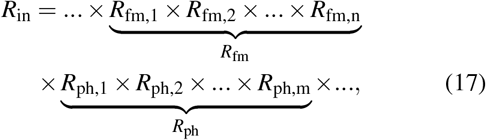

where *R*_fm_ is the infection risk due to the fluid mechanical factors (e.g., ventilation rate, temperature difference and initial velocity of droplets) and *R*_ph_ is the infection risk due to the physiological factors (e.g., the health condition of different people and the survival rate of the virus during transmission). It is important to clarify that in the present study, only *R*_fm_, which is the risk due to the fluid mechanical factors, is analyzed based on the CFD results. The infection risk *R*_fm_ is connected to the simulated aerosol exposure index distributions by an infection model^50^, which is illustrated in Fig. 12. An exposure region in front of a person with volume 2000 ml is defined, and *R*_fm_ is defined by averaging the mean virus-laden aerosol exposure index inside the volume as

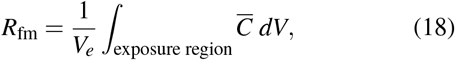

where *V*_*e*_ is the volume of the exposure region.

**FIG. 12.**
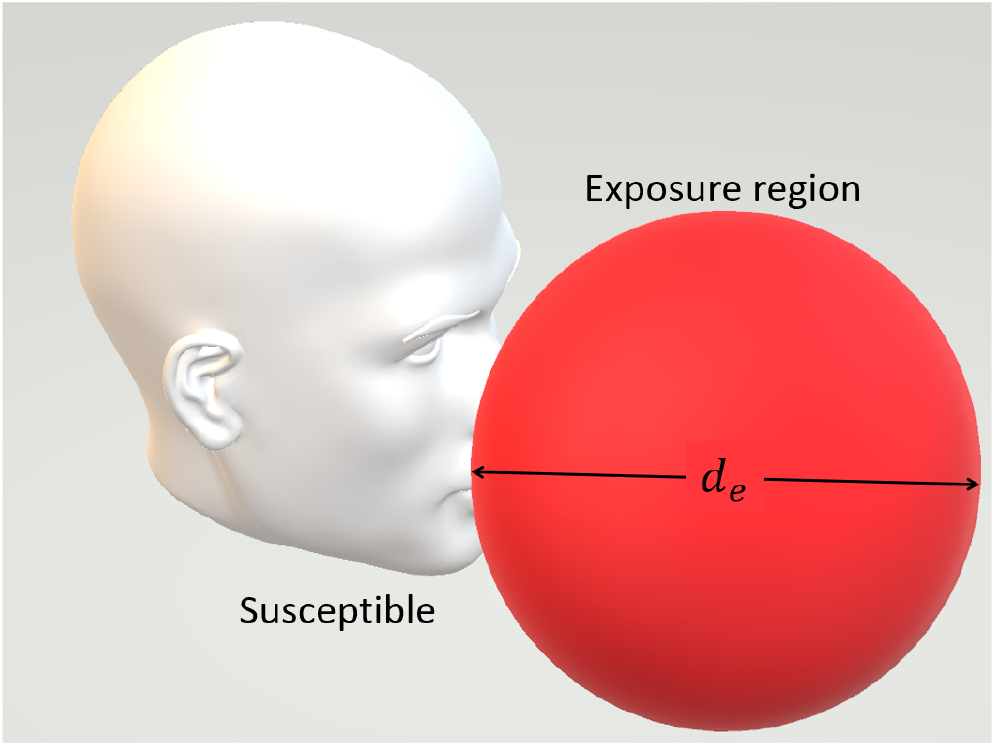
Schematic of a susceptible and his/her exposure region^49^.

The distributions of *R*_fm_ of the customers sitting around T1 are shown in Fig. 13. For the cases without the consideration of thermal effect, it is observed from Figs. 13(a) and (c) that in cases AC14DT00 and AC112DT00, T1P1 has the highest *R*_fm_, T1P3 has the lowest *R*_fm_, and T1P2 and T1P4 are in between. As illustrated by the aerosol exposure index and streamlines shown in Sec. III B, this distribution is due to the recirculation zone above T1, which creates a negative gradient of 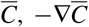, roughly pointing from T1P1 to T1P3. For the cases with the consideration of thermal effect, Fig. 13(c) shows the similar trend in case AC14DT10 compared with the observations from Figs. 13(a) and (b). However, the mechanism that leads to this distribution is different. T1P2 is located in a region where 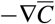 is roughly pointing from T1P1 to T1P2, due to the local thermal plume-formed recirculation zone, while T1P4 is located in a region where 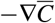 is roughly pointing from T1P1 to T1P4, due to the hot air flows passing by. T1P3 is located the furthest to T1P1. Moreover, comparing Fig. 13(d) with Figs. 13(a), (c), and (d), it is observed that the *R*_fm_ value in case AC112DT10 is around 10% of those in cases AC14DT10 and AC112DT00, and 1% of that in case AC14DT00, because T1P1-T1P4 are all located in the region where aerosols are less concentrated and most of the local air flows are directly from AC1. In the real infection event that occurred in this restaurant, T1P3 is the only person who was not infected during the lunch^9^. Without considering the factors beyond fluid mechanics, the present predictions around T1 in cases AC14DT00, AC14DT10, and AC112DT00 show a same trend as in the real infection.

**FIG. 13.**
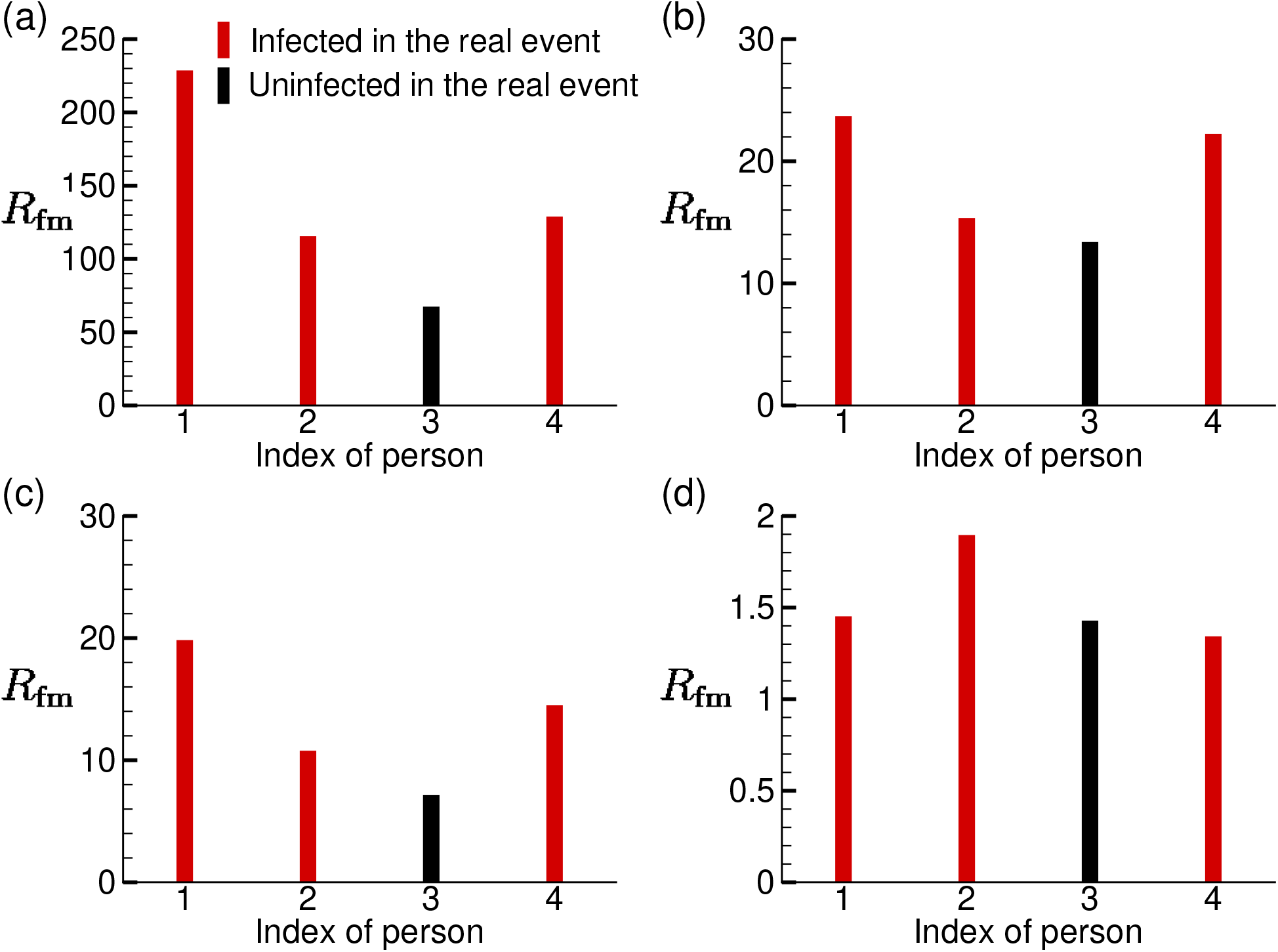
Distributions of the infection risk *R*_fm_ of the customers sitting around T1 in the simulation cases: (a) AC14DT00, (b) AC14DT10, (c) AC112DT00, and (d) AC112DT10.

The *R*_fm_ of the customers sitting around T3 is displayed in Fig. 14. It is observed from Figs. 14(a)-(c) that the distributions of *R*_fm_ in cases AC14DT00, AC14DT10, and AC112DT00 have an approximate V-shape, where the two ends have higher *R*_fm_ than those in between. The difference is that the ratio between the left end in the V-shape distribution to the right is lower in cases AC14DT00 and AC112DT00 than in case AC14DT10. The V-shape distributions observed in cases AC14DT00 and AC112DT00 are due to the air flow sucked into AC1, which creates a distribution of 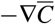 above T3 roughly pointing from T3P7 to T3P4 and from T3P6 to T3P3. However, the V-shape distribution observed in case AC14DT10 is due to a more complex mechanism. Around T3P1 and T3P2, 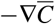 is created by the hot air flows interacting with the cold jet, while around T3P7 and T3P6, the 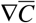 is created by the hot air flow being sucked into AC1. Moreover, a distribution of a different shape is observed in case AC112DT10 in Fig. 14(d), where the *R*_fm_ values of T3P6 and P3P7 are much larger than those of the other five people (the ratios are higher than 2). Similar to the mechanism in case AC14DT10, the distribution of 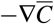 is due to the combined effect of the hot air flows being sucked by AC1 (around T3T7 and T3T6) and the hot air flows interacting with the cold jet (T3P2). The observed much higher risks of T3P6 and T3P7 is due to the stronger cold jet in case AC112DT10 that drives the concentrated region close to the surface of T3. In the real infection event, it is believed that T3P3 and T3P6 were infected during the lunch directly^9^. The prediction on the distributions of *R*_fm_ does not show a higher *R*_fm_ of T3P3 and T3P6 than other people. However, it shows the approximate V-shape distributions in cases AC14DT00, AC112DT00, and AC14DT10, which indicate that the people sitting at the two ends of the table have more risks than the ones in between, which is consistent with the real infection at T3.

**FIG. 14.**
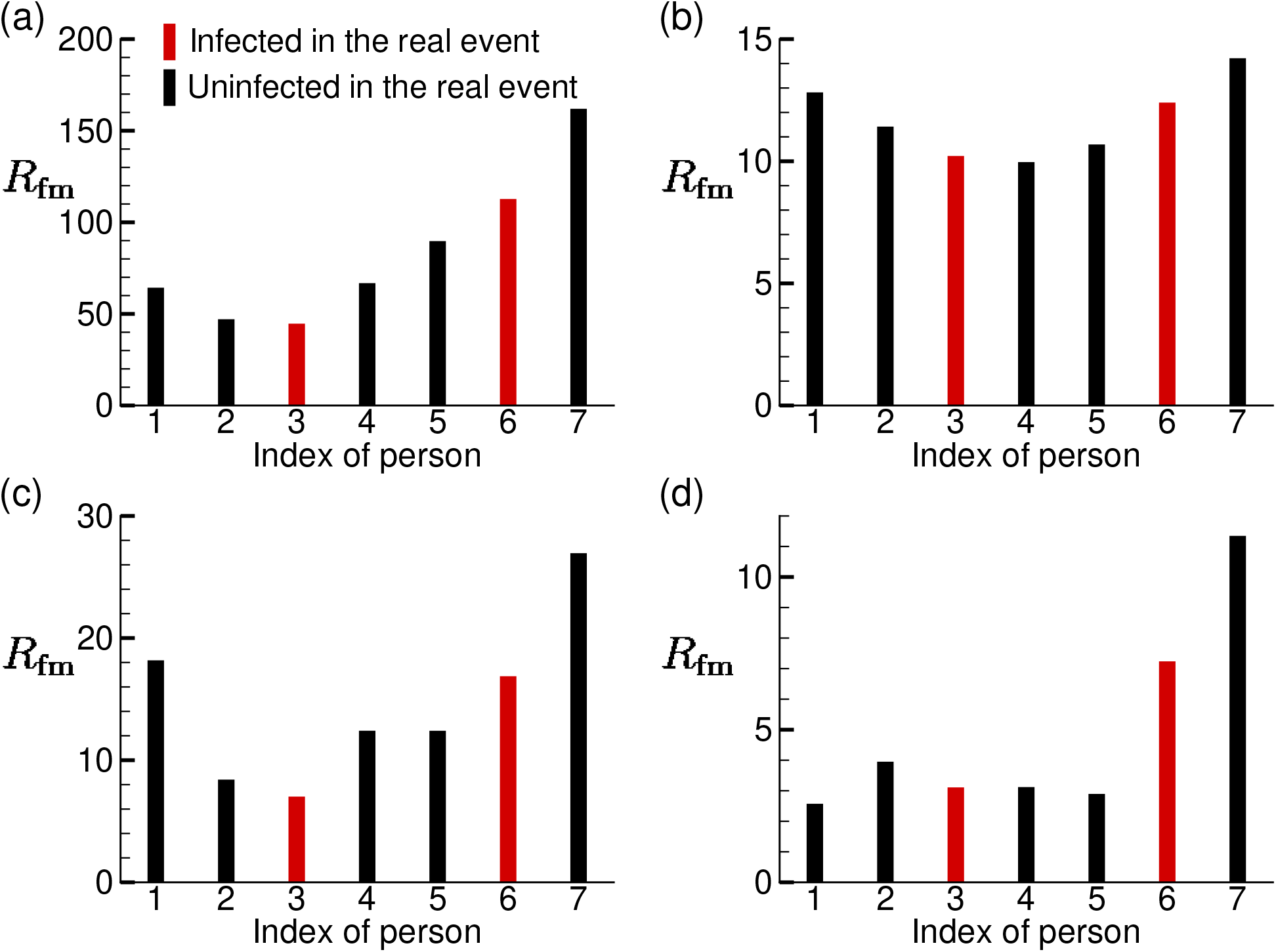
Distributions of the infection risk *R*_fm_ of the customers sitting around T3 in the simulation cases: (a) AC14DT00, (b) AC14DT10, (c) AC112DT00, and (d) AC112DT10.

## D. Further discussions

In this section, further discussions are provided based on the simulated results.

First, we would like to offer some remarks on the applicability of our CFD for infection risk prediction. Specifically, our results have shown that CFD can provide detailed information on the spatial variation of aerosol concentration and aerosol exposure time as well as aerosol transport process to illustrate airborne transmission process and show remarkable agreement with infection pattern reported in the real scenario. Moreover, the CFD results can be used to systematically evaluate the effect of complex environmental factors such as ventilation design, thermal plumes and turbulence on indoor airborne transmission risks. Such information may assist the development of more accurate reduced order models for risk assessment and the implementation of preventive measures under different indoor settings. Nevertheless, we acknowledge that the infection risk evaluation from our current CFD is only derived from the aerosol exposure index. To yield a more substantiated metric of infection risk, a relevant infection-dose model, currently not available for SARS-CoV-2, is needed. In addition, our prediction of aerosol exposure index can be further improved if the more detailed information about the geometry of the space, ventilation, air-conditioning, thermal condition, and human behaviors is accessible.

Second, beyond a quantification of aerosol exposure index, we would like to highlight the ability of our CFD to determine the potential airborne transmission pathways, which can directly lead to actionable preventive measures. Here we use a reverse-time tracing method to determine the origin of aerosols that cause the possible infection exposure of an individual. Specifically, for example, Fig. 15 depicts two intriguing pathways that lead to the infection of T3P3. One, marked by the blue and green aerosols in the figure, shows the potential infection exposure associated with the aerosol transport under the tables. This result suggests the need to shield the space under a table, in addition to the space above, when the interplay between ventilation flow and thermal plumes induces the complex indoor air flows. The other pathway, highlighted with red aerosols, points to the infection potentially caused by the returning aerosols from the AC due to limited filtration efficiency. To further quantify relative significance of such a transmission pathway, we introduce an AC exposure fraction factor, defined as

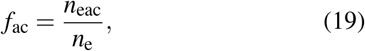

where *n*_e_ and *n*_eac_ are the total number of aerosols and the number of aerosols from AC1 that enter the exposure region during the averaging time. From Fig. 16(a), it is shown that *f*_ac_ can reach as high as 30% for the individuals sitting around T1 in case AC112DT10, due to the directional transport of aerosols through the air flows generated straight from AC1 as shown in Fig. 8(d). For the same reason, in cases AC14DT00 and AC112DT00, T1P3 yields the peak value of *f*_ac_ at about 15%. Among all the cases considered, the lowest *f*_ac_ takes place in AC14DT10 because the air flow from AC1 is strongly disturbed by the rising thermal plumes causing more omni-directional dispersion of aerosols in this case. In contrast, the T3P1 in case AC112DT10 has the maximum *f*_ac_ among all the case, due to the directional transport of aerosols through the jet flow from AC1. Finally, at T3, the *f*_ac_ peaks at T3P3 due to its location adjacent to AC1 with values of about 20% and 11% for the cases AC14DT00 and AC112DT00, respectively. These results indicate that the aerosols returning from ACs is a potentially important factor when evaluating infection risks, and an increase of AC filtration efficiency is highly desirable for mitigating indoor infection risks.

**FIG. 15.**
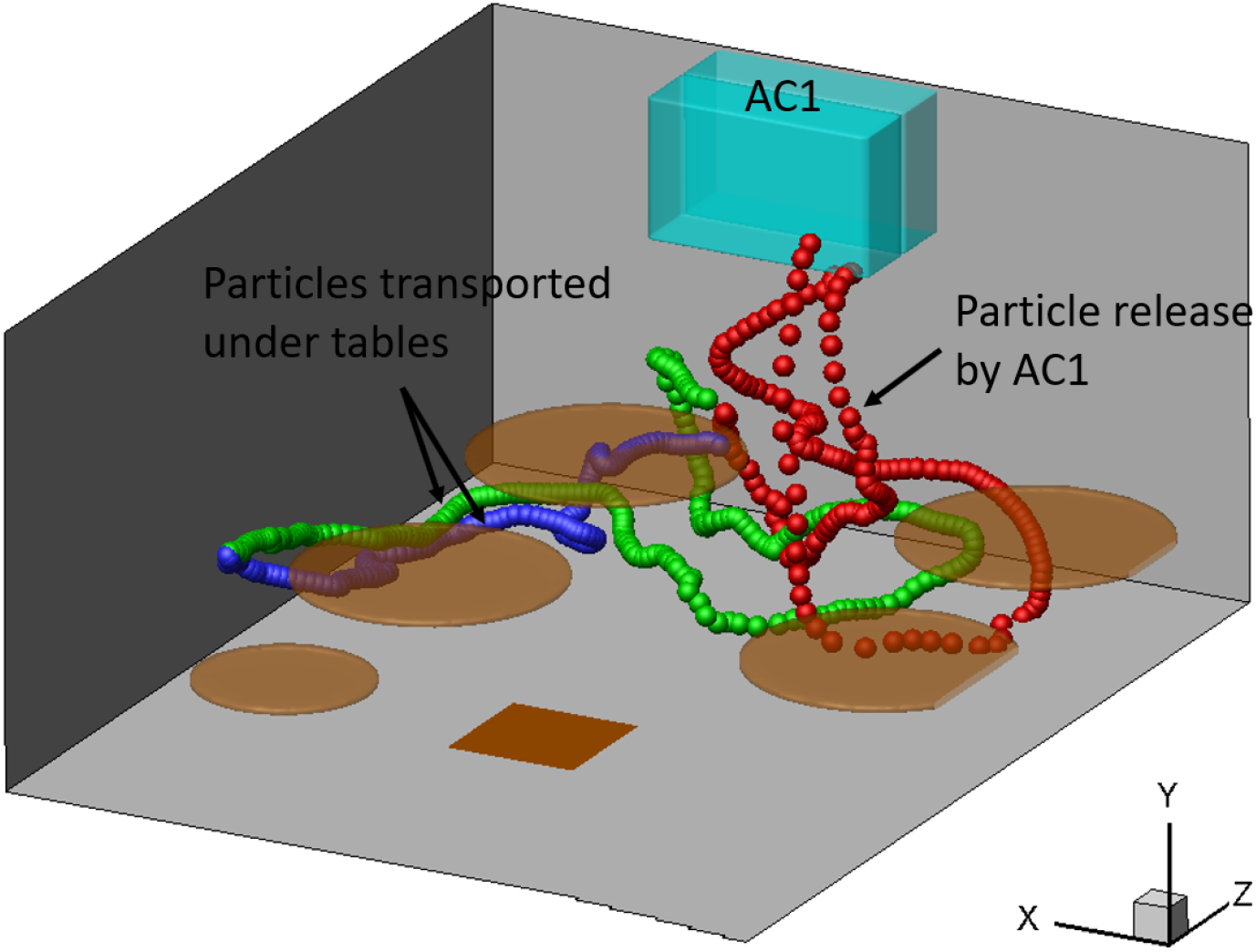
Selected aerosol trajectories traced back from the exposure region of T3P3 to patient zero in case AC112DT10.

**FIG. 16.**
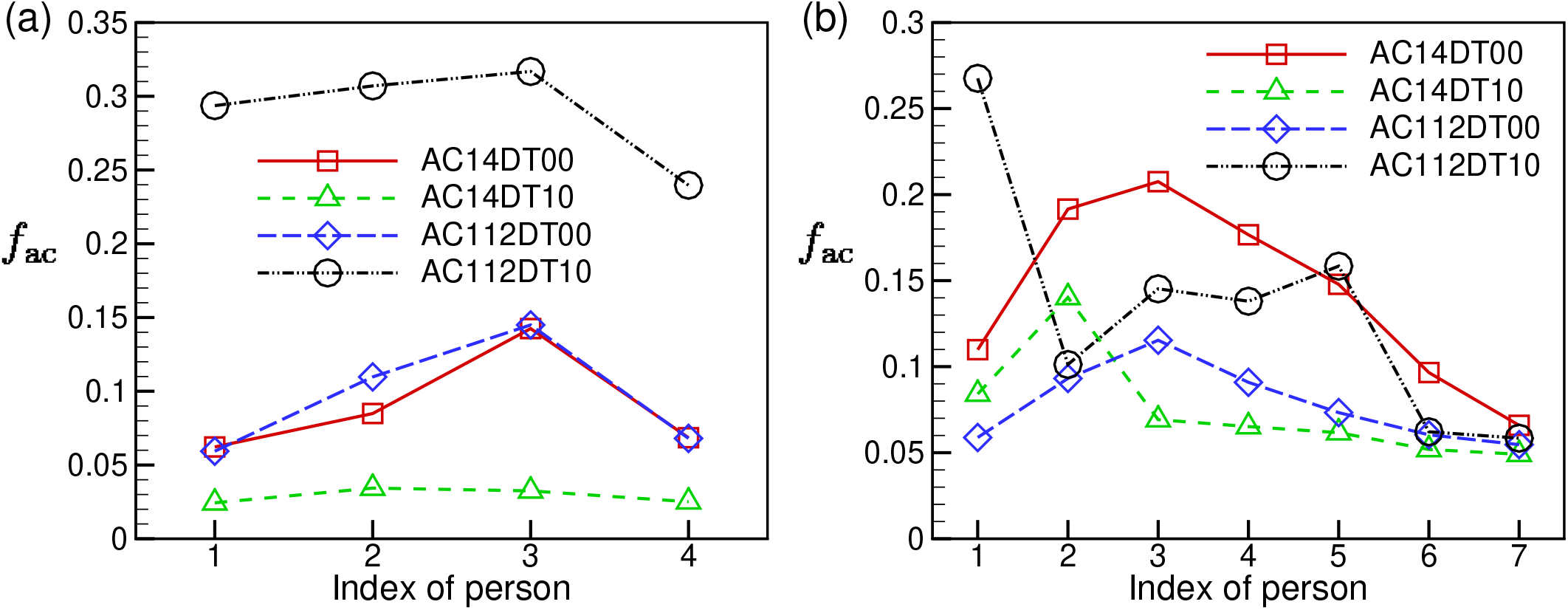
Variations of *f*_ac_ in the exposure regions among the people sitting around (a) T1 and (b) T3.

## IV. SUMMARY AND CONCLUSIONS

In this paper, we presented a systematic CFD-based investigation of indoor airflow and the associated aerosol transport in a restaurant setting, where likely cases of airborne infection of COVID-19 caused by asymptomatic individuals were reported and detailed information of infection process through contact tracing is available. We employed an advanced inhouse LES solver and other cutting-edge numerical methods (i.e., IB method, stochastic modeling of Brownian motion and effect of SGS flow structures on aerosols, Cunningham slip correction, etc.) to resolve complex indoor processes simultaneously, including turbulence, flow–aerosol interplay, thermal effect, and the filtration effect by air conditioners. Using the aerosol exposure index derived from our simulation, we are able to provide a spatial map of the airborne infection risk in the restaurant under different air-conditioning and thermal settings. Our results have shown a remarkable direct linkage between regions of high aerosol exposure index and the reported infection patterns in the restaurant, providing strong support to the airborne transmission occurring in this widely-reported incidence. Moreover, using flow structure analysis based on the mean flow streamlines and the distribution of turbulent kinetic energy and reverse-time tracing of aerosol trajectories in the simulation, we are able to further pinpoint the influence of different environmental parameters (i.e., thermal, ventilation rate, and filtration efficiency) on the infection risks and various potential airborne transmission pathway that can lead to the infection. Specifically, the thermal plumes caused by the temperature difference between ambient air and human/table is shown to cause local recirculation flows in low-ceiling/confined spaces that can substantially increase infection risks. In addition, the interplay between thermal plumes and ventilation flow from air conditioners can result in complex flow patterns that can transport the aerosol circumventing regular shielding and lead to a potential exposure of an individual behind a shield. Moreover, the returning aerosols from air conditioning/ventilation systems due to the limited filtration efficiency can also cause aerosol exposure of individuals adjacent to or facing the ventilation outlets. These results highlight the need for a proper shield design or placement of shielding according to the local flow patterns as well as the need for improving the filtration efficiency of our air conditioning/ventilation system. Overall, our research has demonstrated the capability and value of high-fidelity CFD tools for airborne infection risk assessment and the development of effective preventive measures, which can be further strengthened if proper infection-dose models and more detailed information of specific settings is available.

## Data Availability

The data that support the findings of this study are available from the corresponding author upon reasonable request.

## ACKNOWLEDGMENTS

Jiarong Hong would like to acknowledge the support of University of Minnesota Rapid Response Grant from Office for Vice President of Research (OVPR).

## Appendix A: Validation

Two benchmark cases from the literature were simulated to validate our simulation algorithm. Case 1 is about forced convection, which was measured by Posner *et al*.^51^ The setup of this case is shown in Fig. 17. The dimension of the room model is 0.914 m *×* 0.305 m *×* 0.457 m. A square inlet and a square outlet vents are located on the ceiling of the room and the length of their sides is 0.1 m. The inlet velocity is 0.235 m/s. A partition of height 0.15 m is located at the center of the room. The grid number used in LES is 120 *×* 40 *×* 60. Besides the inlet and outlet boundary conditions at the vents, no-slip boundary condition is applied on the walls and the partition. We compared the measured and simulated vertical velocity along two lines as shown in Fig. 17, where Line 1 is parallel to the central axis of the inlet vent and Line 2 goes through the center of the partition. The comparison is plotted in Fig. 18. As shown, the present LES result is very close to the experiment result.

**FIG. 17.**
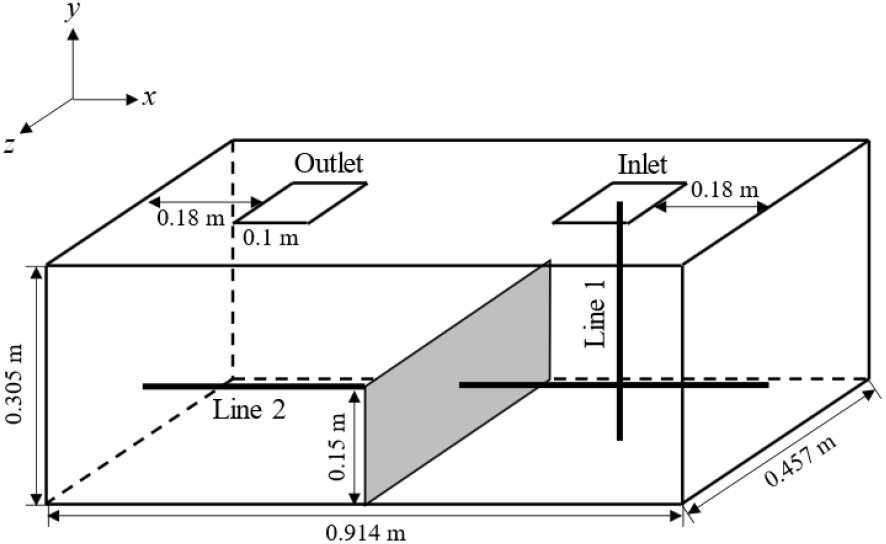
Set-up of validation Case 1.

**FIG. 18.**
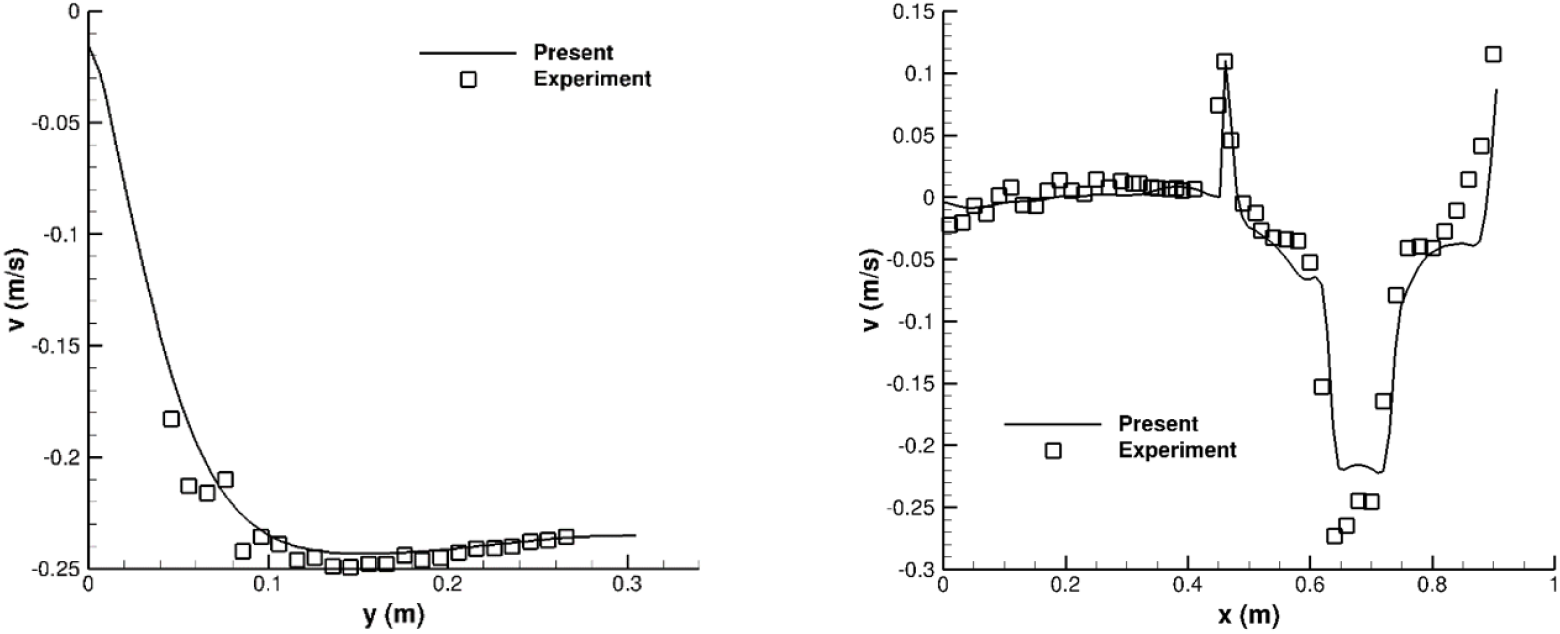
Comparison of vertical velocity along (a) Line 1 and (b) Line 2 in Fig. 17 from the present LES and the previous experiment^51^ for the validation Case 1. Lines 1 and 2 are marked in Fig. 17.

We have also conducted convergence tests on our numerical algorithm. We performed LES for Case 1 with the grid numbers of 80 *×* 30 *×* 45, 120 *×* 40 *×* 60, and 180 *×* 60 *×* 90. The comparison of the vertical velocity along Line 1 is plotted in Fig. 21. As shown, our simulation algorithm obtains good convergence performance with different grid resolutions.

Case 2 is about mixed convection, which was measured by Blay *et al*.^52^ The set-up of this case is shown in Fig. 19. The dimension of room model is 1.04 m *×* 1.04 m *×* 0.7 m. An inlet vent and an outlet vent span along the *z*–direction, and their widths are 0.018 m and 0.024 m, respectively. The inlet velocity is 0.57 m/s. The grid number is 120 *×* 120 *×* 60. The boundary conditions for the flow velocity are the same as those in Case 1. For the set-up of heat transfer, the floor temperature is 35 °C, the temperature on all the walls and the ceiling is 15 °C, and the temperature of the inlet flow is also 15 °C. We compared our velocity and temperature results along Line 1 (*x* = 0.52 m, *z* = 0.35 m) and Line 2 (*y* = 0.52 m, *z* = 0.35 m) with the previous experiment^52^. As shown in Fig. 20, our simulation agrees with the measurement. Grid convergence test for validation Case 2 have also be performed in a way similar to validation Case 1 and is not repeated here.

**FIG. 19.**
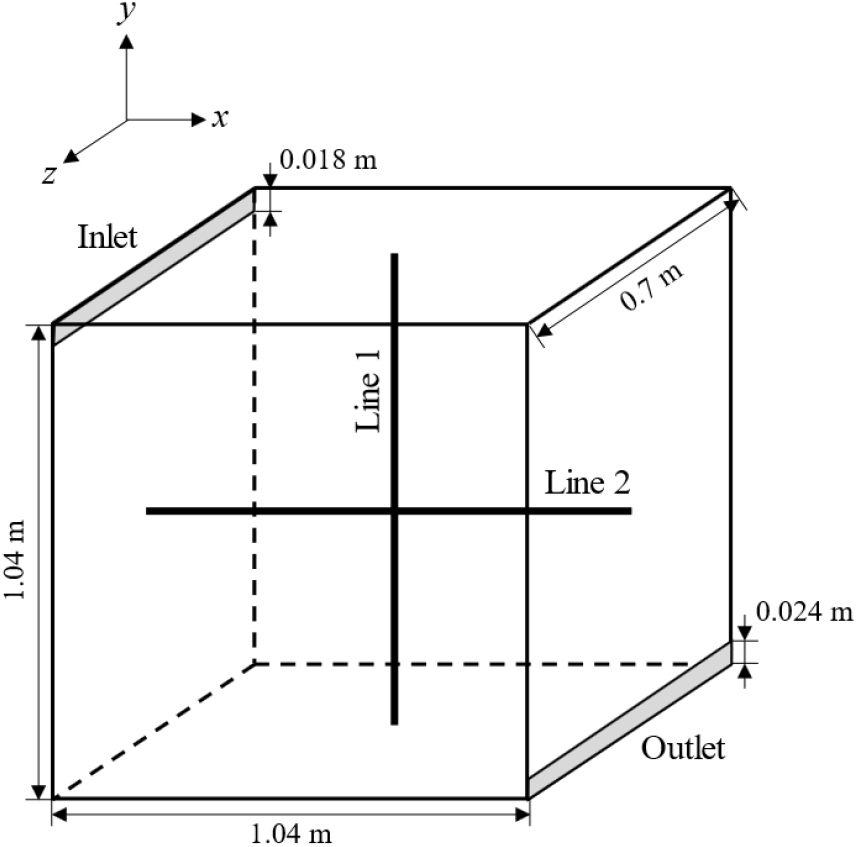
Set-up of validation Case 2.

**FIG. 20.**
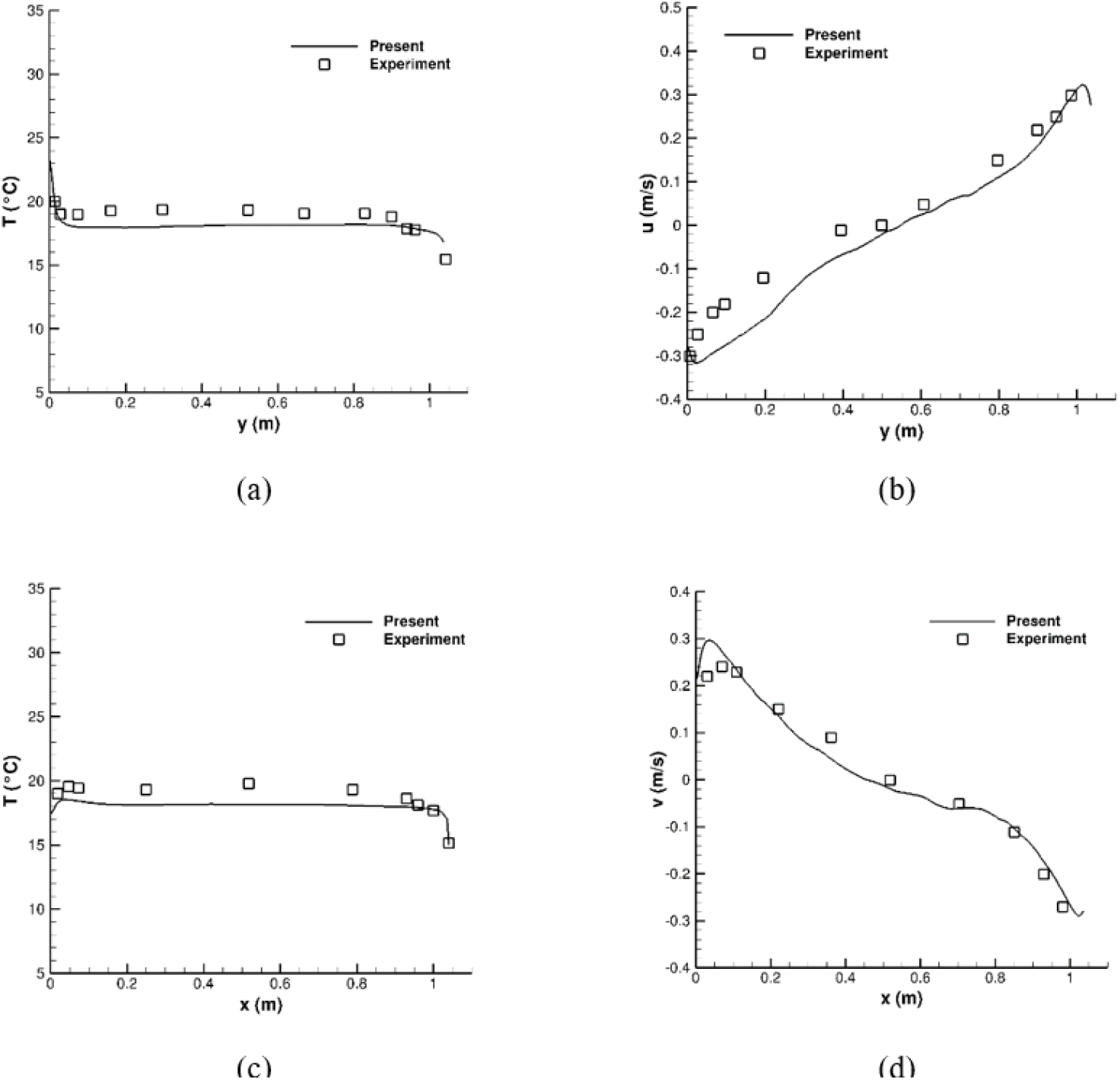
Comparison of the results for validation Case 2 from the present LES and the previous experiment^52^. (a) Temperature along Line 1, (b) horizontal velocity along Line 1, (c) temperature along Line 2, and (d) vertical velocity along Line 2. Lines 1 and 2 are marked in Fig. 19.

**FIG. 21.**
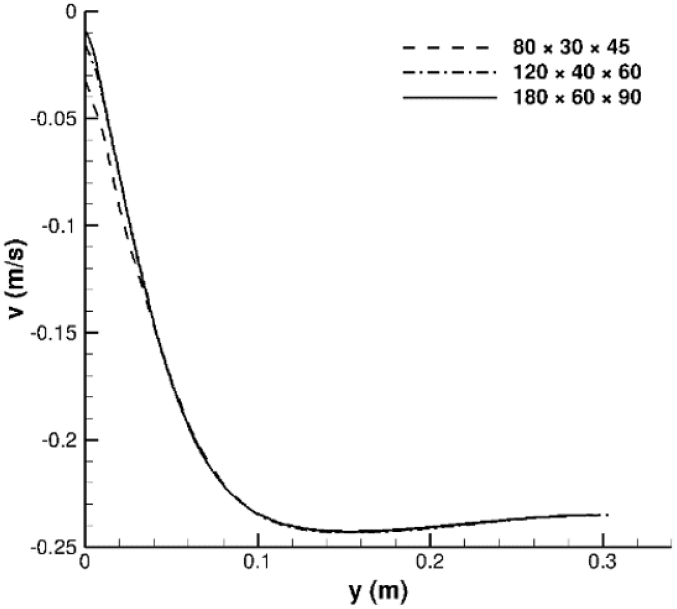
Grid convergence test of the vertical velocity along Line 1 in Case 1.

